# Early Mucosal Type II Interferon Limits SARS-CoV-2 Replication in Humans

**DOI:** 10.64898/2026.06.29.26356894

**Authors:** Alexander Viloria Winnett, Alexandra Tabachnikova, Jonathan Chen, Kerrie Greene, Anna E Romano, Xinyue Penny Pei, Matthew M Cooper, Julio Silva, Alyssa M Carter, Jialong Jiang, Yong Kong, Morgan Roos, Christina Middle, Hanqiao Zhang, Matt Thomson, Keith Booher, Scott Kuersten, Akiko Iwasaki, Rustem F Ismagilov

## Abstract

COVID-19 vaccines markedly reduce disease severity, but their ability to block infection and transmission remains limited and variable.^1^ A better understanding of early mucosal immune programs that constrain viral replication at susceptible upper respiratory sites is needed to develop more effective antiviral strategies. However, temporal and anatomical antiviral dynamics are difficult to resolve without longitudinal, paired-site sampling beginning before infection onset. We quantified longitudinal viral load by RT-qPCR and human gene expression by mRNA sequencing in 1,237 samples prospectively collected daily from the nasal cavity, oral cavity, and oropharynx of 16 individuals starting from the onset of naturally acquired SARS-CoV-2 infection, and 16 age-, sex-, and vaccination-matched uninfected individuals. Here we show that Type I interferon (IFN) responses are initiated concurrently across these upper respiratory sites, even before local viral detection. In contrast, Type II IFN initiation is more spatially variable, and earlier nasal Type II IFN initiation is associated with reduced viral replication, prior COVID-19 vaccination, and higher tissue-resident memory T cell (T_RM_) signature expression. These findings demonstrate that in addition to humoral immunity, prior vaccination primes rapid, inducible mucosal Type II IFN responses, likely mediated by rare T_RM_ upon viral encounter, to limit viral replication and spread.

**ONE SENTENCE SUMMARY:** Using a unique study design with temporally-dense, paired sampling of the oral cavity, nasal cavity and oropharynx from the onset of naturally acquired human SARS-CoV-2 infection, we demonstrate that Type I IFN responses are initiated synchronously in the upper respiratory mucosa during early infection, whereas the timing of Type II IFN response initiation is asynchronous but earlier initiation is associated with prior COVID-19 vaccination, enhanced tissue-resident memory T cells signatures, and reduced local viral replication.

**Graphical Abstract.:** Among participants who prospectively collected daily, paired specimens from multiple upper respiratory anatomical sites while enrolled in a case-ascertained COVID-19 household transmission study conducted in Los Angeles, California between September 2020 and April 2022, a subset of participants were found to initially be negative upon enrollment but later became positive for SARS-CoV-2. These were defined as Cases of naturally acquired, incident SARS-CoV-2 infection, and for whom age, sex, and prior COVID-19 vaccination status matched Control participants without SARS-CoV-2 infection were identified, and demographic data is shown. Samples underwent viral load quantification by RT-qPCR, validated by RT-ddPCR, and human transcriptome sequencing to calculate immune pathway activation and determine the timing of Type I and II IFN response initiation, and expression of a tissue resident memory T cell (T_RM_) signature. Differences in the magnitude and timing of these pathways were compared by vaccination status, and by subsequent viral RNA shedding to identify mucosal immune correlates of reduced viral replication. Created in https://BioRender.com.

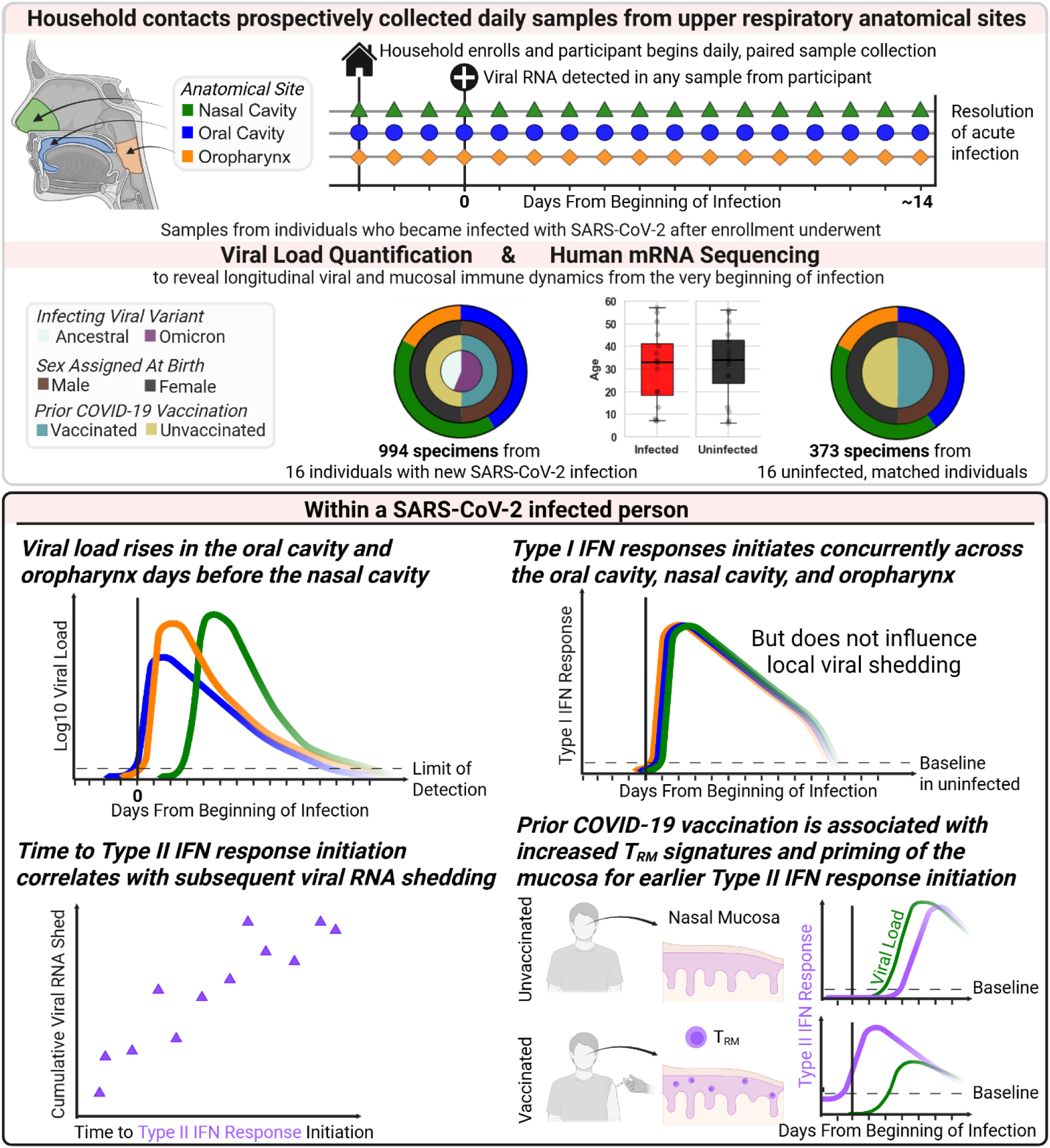

## INTRODUCTION

Current COVID-19 vaccines provide robust protection against severe disease and can attenuate breakthrough infection, but do not completely prevent onward transmission to new hosts.^1,2^ Transmission sustains opportunities for the emergence of variants that escape immunity conferred by prior vaccination or infection.^3^ Vaccines that prevent onward transmission can interrupt this cycle to reduce infection prevalence and the necessary scale of downstream infection control measures.^4^ Understanding how early immune responses control viral replication at the primary site of infection, the upper respiratory mucosa,^5^ is necessary to develop vaccines that limit breakthrough infections, viral shedding and onward transmission.^4^

While considerable work has focused on optimizing antibody-mediated protection following COVID-19 vaccination,^6^ early local mucosal immune responses that constrain viral replication including interferon (IFN) responses have been relatively underinvestigated.^7^ The synchronization of IFN response initiation among upper respiratory sites (e.g., nasal cavity, oral cavity, oropharynx, nasopharynx) within an individual during early SARS-CoV-2 is unknown. Additionally, how the timing of initiation for different IFN types (Type I, II, and III) at the onset of naturally acquired SARS-CoV-2 infection in the upper respiratory mucosa of humans influences local viral replication has not been demonstrated. It is also unclear whether prior COVID-19 vaccination modulates the timing of IFN response initiation in the human upper respiratory mucosa during breakthrough infection.

Available evidence suggests that the timing of IFN initiation is a key determinant of early viral control. Type I (IFN-α and IFN-β) and Type III IFNs (IFN-λ) are mostly produced by infected cells after sensing viral pathogen-associated molecular patterns (PAMP), and signal through interferon-stimulated gene factor 3 (ISGF3) complexes to elicit partially overlapping downstream interferon-stimulated genes (ISGs), with Type III IFN responsiveness enriched at epithelial barriers.^8^ Type II IFN (IFN-γ) is secreted by lymphocytes, such as natural killer cells, innate-like lymphocytes and T cells, including tissue resident memory T cells (T_RM_),^9^ and links innate PAMP-driven activation with antigen-specific adaptive immunity.^10^ In mice, early Type I and III IFN responses abrogate SARS-CoV replication,^11^ but permit greater MERS-CoV replication when delayed.^12^ Early induction of Type II IFN reduced peak SARS-CoV-2 viral loads in the lungs of mice,^13^ and in a human household transmission cohort, the abundance of SARS-CoV-2-specific IFN-γ-secreting T cells one week after exposure anticorrelated with peak viral load.^14^

Obtaining longitudinal measurements of distinct IFN responses in upper respiratory mucosal surfaces from the onset of SARS-CoV-2 infection in humans is logistically and technically challenging. High frequency sample collection beginning before or at the onset of infection is necessary to characterize how early upper respiratory mucosal immune responses shape subsequent viral replication. Such samples have been obtained in controlled human viral challenge studies, which typically involve experimentally inoculating a small cohort (<20 individuals) of young, healthy participants,^15,16^ as well as prospective cohort studies and case-ascertained household transmission studies,^14,17^ though the onset of infection is observed in only a small fraction of participants and sampling is typically less frequent or from a single upper respiratory site.

To investigate how the synchronization and timing of distinct early IFN responses in the upper respiratory mucosa impact SARS-CoV-2 replication, we conducted a case-ascertained household transmission study in the Los Angeles area between 2020-2022. Among over 400 participants, 16 were SARS-CoV-2–negative at enrollment but subsequently developed sustained incident infection. These participants, together with 16 age-, sex-, and vaccination-matched uninfected participants, collected daily or twice-daily samples from the nasal cavity, oral cavity, and oropharynx for approximately two weeks. From these samples, we quantified longitudinal viral loads,^18,19^ and performed bulk mRNA sequencing to characterize the synchronization and timing of IFN responses among upper respiratory mucosal sites from the onset of infection, evaluate how the timing of IFN initiation impacts subsequent viral replication, and determine whether prior COVID-19 vaccination modulates early mucosal IFN dynamics. Together, our results reveal the timing of Type II IFN response as a modulator of local SARS-CoV-2 replication, and a potential correlate of protection for vaccine candidates intended to block onward transmission.

## RESULTS

To interrogate early mucosal immune dynamics, we conducted a case-control study of 16 participants aged 7 to 57 years old with incident SARS-CoV-2 infection, as well as 16 age-, sex-, and COVID-19 vaccination-matched uninfected controls (**Table S1**). We optimized methods to overcome technical challenges inherent to self-collected human upper respiratory specimens, then quantified longitudinal viral loads and performed human exome-enriched RNA sequencing on 1,273 specimens from paired upper respiratory mucosal sites (oral cavity, nasal cavity, oropharynx) collected daily or twice daily by each participant (**Figure 1, Figure S1**). Following RNA extraction, RT-qPCR analysis of a human housekeeping gene, *MYH9*, revealed only minor sampling variation over time within each participant (**Figure S2A-C**). Successful cDNA libraries were generated from 99% of all self-collected upper respiratory human clinical specimens attempted (1262 of 1273, **Figure S2D**). Despite high microbial background, human exome enrichment enabled greater than 10 million human mapping reads in over 90% of samples attempted (1165 of 1273, **Figure S2E**) with high reproducibility among technical replicates (**Figure S2F-H**). Gene expression data exhibited anticipated biological patterns by participant sex and anatomical sampling site (**Figure S2I,J)**. From this unique dataset, we investigated the relative timing and synchronization of IFN responses among upper respiratory sites within SARS-CoV-2 infected individuals, how early IFN responses relate to viral loads throughout infection, and the impact of prior COVID-19 vaccination.

**Figure 1.**
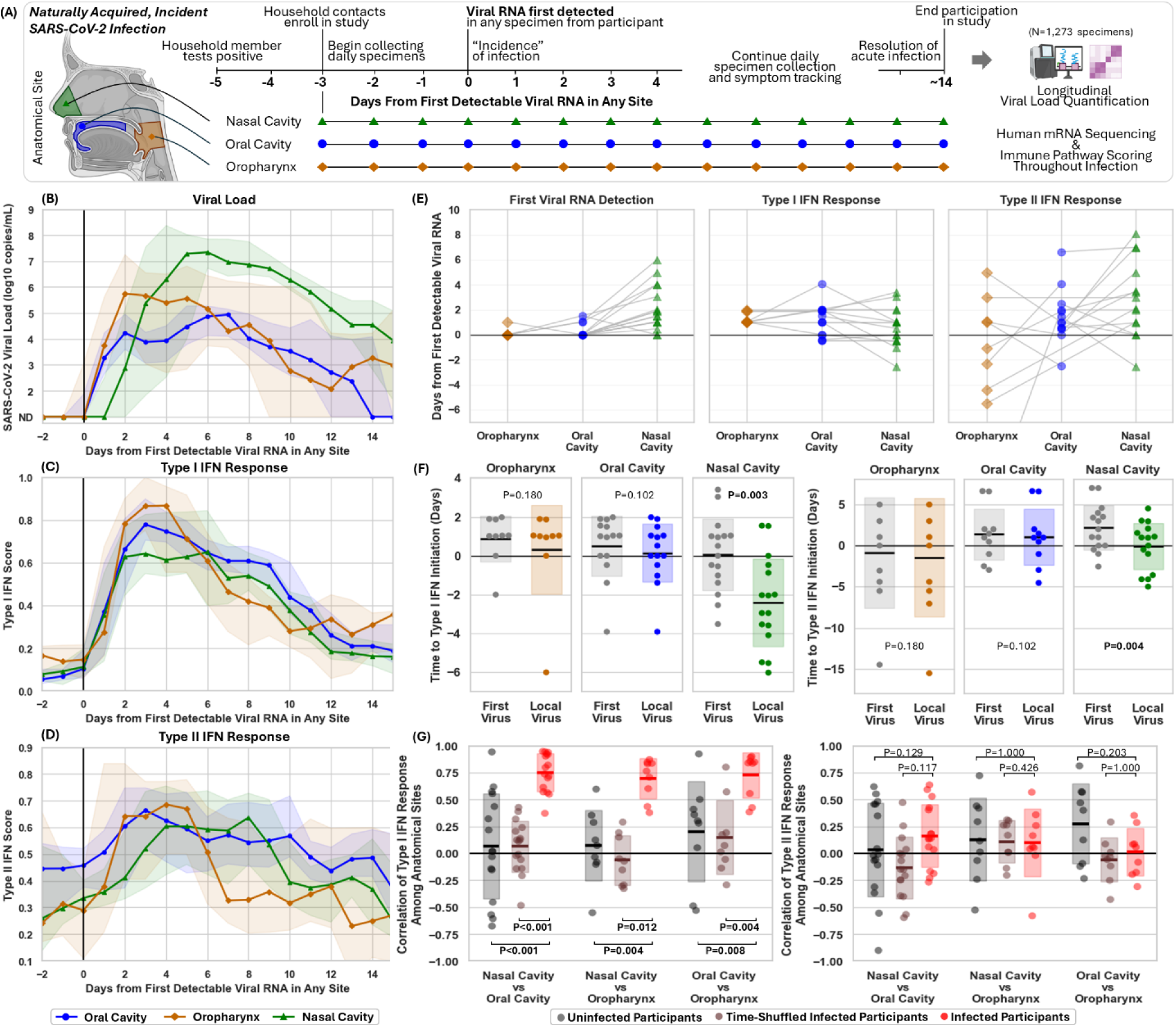
The Type I IFN response is initiated concurrently despite extreme differences in early local viral loads among upper respiratory sites within an individual, the whereas the Type II IFN response is not. (A) Prospective, daily collection of specimens from multiple upper respiratory sites enables profiling of longitudinal host-pathogen dynamics from the very beginning of naturally acquired SARS-CoV-2 infection. (B) Median viral load and bootstrapped 95% CIs were plotted by day relative to first SARS-CoV-2 detection at any anatomical site, stratified by site. (C–D) Mean Type I and Type II IFN module scores were calculated in 2-day windows, participant-normalized within site, and plotted with bootstrapped 95% CIs. (E) Within-participant timing of first site-specific viral detection >10³ copies/mL, Type I IFN initiation, and Type II IFN initiation is shown across anatomical sites, with lines connecting sites from the same participant. (F) For each site, the “Local Virus” interval was defined as time from IFN response initiation to viral load >10³ copies/mL at the same site, and the “First Virus” interval as time from IFN response initiation to viral load >10³ copies/mL at any site. Points represent participants; intervals were compared by Wilcoxon signed-rank test with Bonferroni correction. (G) IFN coordination was assessed by within-participant correlations of IFN module scores across anatomical sites in infected participants and compared with two null distributions: correlations across sites in uninfected participants and correlations after random shuffling of one infected-participant time series. Distributions were compared by Wilcoxon rank-sum test with Bonferroni correction; infected-participant median daily IFN module scores with bootstrapped 95% CIs are shown by site.

### Early Type I IFN responses were synchronized across the upper respiratory mucosa of individuals infected with SARS-CoV-2, and often preceded viral detection in the nasal cavity

Since SARS-CoV-2 can infect multiple anatomical sites within the human upper respiratory mucosa,^20^ early mucosal immune responses at each site and their coordination can influence the course of infection. Therefore, we first sought to investigate the relationship between timing of viral presentation and IFN responses.

Robust and well-validated gene sets were available for Type I and Type II IFN responses, but not for Type III IFN responses. Published Type III IFN gene sets (GO:0034342^21^ and M39464^22^) were small (<15 genes) and dominated by *IFNL* transcripts, which at the bulk tissue level were lowly expressed, resulting in gene set module scores that poorly differentiated infected from uninfected participants (**Figure S3**).

Within infected individuals, SARS-CoV-2 presents in the nasal cavity several days after the oral cavity or oropharynx.^18,19^ Among infected participants, detection of SARS-CoV-2 in the nasal cavity occurred at least one day later than the oral cavity or oropharynx for 88% of participants (**Figure S1**), on average, 2.3 days later. Aggregated trajectories demonstrate this delayed rise in nasal cavity viral loads relative to the oral cavity and oropharynx (**Figure 1A**). Given these differences in the timing of viral detection among upper respiratory mucosal sites, we asked whether IFN responses were also asynchronous and followed local viral detection.

To our surprise, we found that initiation of the Type I IFN response did not always follow the presence of viral RNA within a site, but initiated concurrently in the oropharynx, oral and nasal cavities shortly after SARS-CoV-2 detection in any site. Canonical Type I IFN-inducible genes (e.g., *STAT2, MX1, MX2, OAS1, OAS2, IFIT1*) exhibited low and stable expression among uninfected participants, but dynamic rises in all upper respiratory anatomical sites from individuals with infection (**Figure S4**), which appeared grossly synchronized across sites within individuals (**Figure S5A,B**). Canonical Type I IFN genes were significantly upregulated in the nasal cavity even before viral RNA was detected in the nasal cavity (**Figure S5C-G**). Module scores based on a predominately Type I IFN response gene set^15,23^ demonstrated synchronization of the Type I IFN response across the upper respiratory mucosa of infected individuals (**Figure S5H,I**).

Type I IFN response module scores aggregated across infected participants exhibited striking synchronization among anatomical sites, with upregulation at all sites immediately after viral detection at any site (**Figure 1B).** We defined the initiation of the Type I IFN response in each participant and anatomical site as the first time point at which a module score exceeded a threshold that optimally distinguished infected from uninfected participants at that site (**Figure S6**). For 81% infected individuals (13 of 16, all except **D, L** and **M** panels in **Figure S7**), initiation of the Type I IFN response in the nasal cavity occurred prior to nasal viral load reaching levels detectable by most high-analytical-sensitivity COVID-19 tests (1000 copies/mL). While detection of viral RNA in the nasal cavity was clearly delayed relative to the oral cavity or oropharynx (**Figure 1A**), initiation of the Type I IFN response did not exhibit this delay (**Figure 1B,D**). We compared the interval between Type I IFN response initiation and viral detection at the same site with that at any site. In the oral cavity and oropharynx, where viral RNA was first detected in most participants, there was no significant difference between these intervals. In contrast, nasal cavity Type I IFN initiation occurred more closely to the first viral detection in any site than local viral detection (**Figure 1E**).

Longitudinal Type I IFN module scores were temporally coordinated across anatomical sites within individuals, independent of viral load trajectories. Type I IFN module scores were highly correlated among anatomical sites within each infected participant (P_adj_<0.012, **Figure 1F**) and did not correlate better with viral load in the same anatomical site than viral load in other sites (**Figure S5I**). Correlation of Type I IFN module scores between pairs of anatomical sites (e.g. nasal cavity and oral cavity in the same individual) was compared to two null models: correlation of Type I IFN module scores from uninfected participants, and from infected participants but with timepoints for one site randomly shuffled. Type I IFN module scores correlated significantly better than these null models for all site-pairs (**Figure 1F**), supporting that Type I IFN responses were strongly correlated across the upper respiratory mucosa of an individual regardless of local viral load. These results were corroborated in two orthogonal analyses: an alternate Type I IFN gene set (**Figure S8**) and single-sample gene set enrichment scores demonstrating activation of viral sensing pathways upstream of Type I IFN responses, such as RIG-I and TLR7 (**Figure S9**). Cell type deconvolution using publicly available scRNA-seq data suggested that conventional dendritic cells (DC), macrophages, and plasmacytoid DCs most likely contribute to the synchronization of the Type I IFN response among the upper respiratory mucosa (**Figure S10**). Notably, CD8 T cells abundance was anti-correlated with type I IFN response signature across the mucosal sites (**Figure S10**).

### Type II IFN response timing was variable across upper respiratory mucosal sites

While concurrent initiation of the Type I IFN response was observed among upper respiratory mucosal anatomic sites within infected individuals regardless of local viral detection, this concurrence was not observed for the Type II IFN response. Module scores for a Type II IFN response gene set (GO:0034341^21^) differentiated infected from uninfected participants (**Figure S11**) and demonstrated dynamic rises during infection (**Figure S12**). However, in contrast to Type I IFN (**Figure 1B**), Type II IFN responses did not exhibit synchronization between anatomical sites during early infection (**Figure 1C**). Rather, initiation of the Type II IFN response appeared more variable among anatomical sites than initiation of the Type I IFN response or detection of virus (**Figure 1D**) and was more closely related to detection of virus within the same site than the Type I IFN response (**Figure 1E**). Type II IFN module scores did not correlate significantly better across anatomic sites within infected individuals than two null models – correlation among time-shuffled data from infected individuals, and correlation among anatomic sites from uninfected individuals (**Figure 1F**). Further, correlation of Type I IFN module scores among site-pairs was markedly higher than the correlation of Type II IFN module scores among site-pairs (**Figure S13**). These results were recapitulated with an alternate Type II IFN gene set (**Figure S14**).

### Early initiation of Type II, but not Type I, IFN response was associated with reduced viral replication

Understanding which immune responses effectively control viral replication and reduce shedding in humans is crucial to designing vaccines that prevent breakthrough infections capable of forward transmission. To investigate this, we next asked whether the timing of Type I or Type II IFN responses within an upper respiratory site impacted subsequent viral replication from that site. We used peak viral load (in RNA genomic copy equivalents per mL of sample) and the integration of RNA viral load measurements over time (cumulative viral load) as indicators of viral RNA shedding.

We analyzed longitudinal Type I and Type II IFN response module scores and viral loads within the nasal and oral cavities of infected individuals. Type I and Type II IFN response module scores exhibited distinct trajectories (**Figure 2A,B**). For each participant, we classified whether the IFN response was initiated early (prior to a viral load of 10^3^ copies/mL within that anatomical site) and tested whether early initiation was associated with reduced viral replication (**Figure 2C,D**). In both the nasal and oral cavities, early initiation of a Type II IFN response was significantly associated with lower cumulative virus shed at day 10 of infection, by one to two orders of magnitude, whereas early initiation of a Type I IFN response was not. Similar results were observed for peak viral load, which was highly correlated with cumulative virus shed by day 10 of infection (**Figure S15A,B**). Of note, most participants were classified as having early initiation of the nasal Type I IFN response because the initiation of this response was synchronized with the earlier presentation of virus in the oral cavity or oropharynx, rather than the later presentation of virus in the nasal cavity (**Figure 1**). One infected participant did not exceed a viral load of 10^3^ copies/mL in the oral cavity and did not reach a module score above the threshold considered initiation of the Type II IFN response. Five other infected participants did not exceed the module score threshold for Type II IFN response initiation; when these participants were classified as having late initiation of the Type II IFN response, earlier initiation still demonstrated significantly reduced viral replication (P_adj_=0.009). The oropharynx exhibited a similar association between early Type II IFN responses and reduced viral replication, though the difference was not statistically significant (**Figure S15C,D**). Oropharyngeal sampling was introduced later in the study than nasal and oral cavity sampling; thus, fewer participants collected samples from this site, and all participants who collected oropharyngeal samples were vaccinated. Aggregated, longitudinal Type II IFN scores appeared to rise earlier among participants who previously received COVID-19 vaccination than those who had not (**Figure 2A,B**).

**Figure 2.**
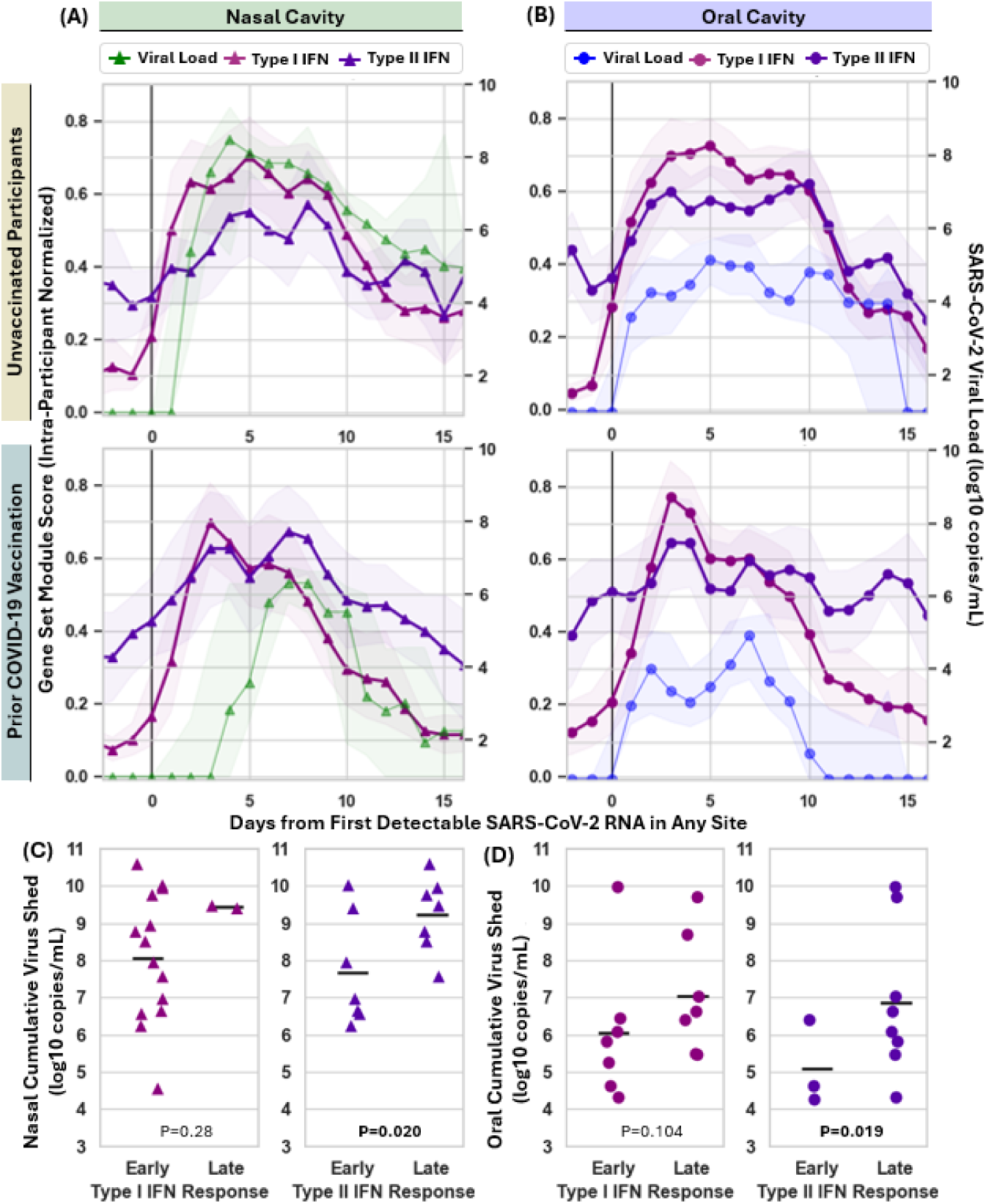
Earlier initiation of Type II interferon responses are associated with reduced viral replication from the nasal and oral cavities of individuals with acute SARS-CoV-2 infection. (A,B) Longitudinal SARS-CoV-2 viral loads (2-day median among infected participants), overlayed with longitudinal Type I and Type II IFN response module scores (2-day mean among infected participants). Data is stratified by individuals who had never received a COVID-19 vaccination and those who had previously received at least one dose of COVID-19 vaccination, and by data from the nasal cavity **(A)** and the oral cavity **(B)**. Shaded regions indicate 95% Confidence Intervals calculated by bootstrapping. Black vertical line indicates day 0 of infection, when viral RNA was detected in any anatomical site within a participant. **(C-D)** Participants were classified as exhibiting early versus late initiation of Type I (left) or Type II (right) IFN responses in the nasal cavity **(C)** or the oral cavity **(D)**. Individuals with early initiation achieved a module score differentiable from uninfected participants prior to a viral load of 10^3^ copies/mL in the nasal cavity. The cumulative virus shed by day 10 of infection for each participant was calculated as the area under curve of daily viral load measurements for that participant. Distributions of cumulative virus shed by day 10 of infection were compared between participants with early versus late initiation of Type I IFN responses (left) and Type II IFN responses (right) by Wilcoxon Rank Sum Test. P-values were adjusted for multiple hypothesis testing using the Benjamini Hochberg method.

### Pre-infection COVID-19 vaccination was associated with earlier Type II IFN responses and *reduced viral replication in the nasal cavity*

We next asked whether prior COVID-19 vaccination was associated with earlier initiation of IFN responses and reduced viral replication. Given that Type I IFN signaling is an innate antiviral response, prior vaccination was neither expected nor observed to associate with earlier Type I IFN response initiation in either the nasal or oral cavities (**Figure 3A,B**).

**Figure 3.**
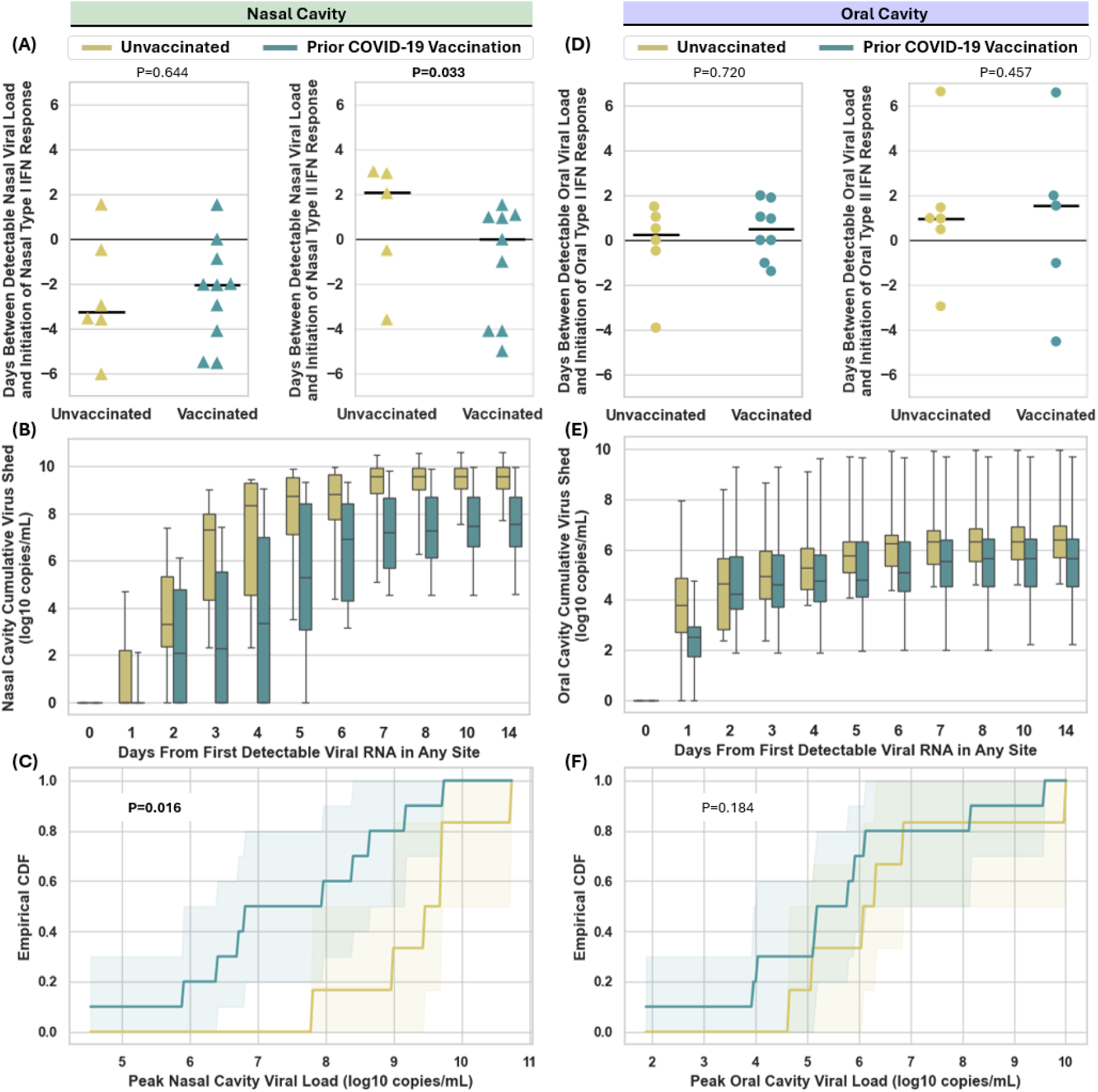
Pre-infection COVID-19 vaccination results in earlier nasal Type II interferon responses and reduced viral replication. After defining a Type I and Type II Interferon Response module score that differentiated between infected and uninfected participants (**Figure S6, Figure S11**), the first timepoint with a module score above this threshold was considered the initiation of that IFN response in the nasal **(A)** and oral cavity **(D)** of each infected participant. For each participant, the time difference between the initiation of the interferon response in that cavity, and the first timepoint with a viral load above 1000 copies/mL in the nasal cavity was calculated and plotted stratified by vaccination status. Mann Whitney U test was performed to assess for a difference in the relative timing of Type II IFN response by vaccination status. For each day of infection, the distribution of cumulative viral loads in the nasal cavity **(B)** and oral cavity **(E)** among vaccinated (aqua) and unvaccinated (gold) infected participants was plotted. Empirical Cumulative Density Functions for peak viral loads in the nasal cavity **(C)** and oral cavity **(F)** of infected individuals were plotted, stratified by those who had received at least one intramuscular COVID-19 vaccine dose (aqua) and those who had not (gold). Mann Whitney U test was performed to test for differences in the distribution of peak viral loads between groups. P-values were adjusted for multiple hypothesis testing using the Benjamini Hochberg method.

In contrast, prior COVID-19 vaccination was associated with earlier initiation of a Type II IFN response in the nasal cavity (**Figure 3A**). Orthogonal analysis using a time-to-event framework further demonstrated significantly earlier initiation of Type II but not Type I IFN responses in the nasal cavities of vaccinated individuals (**Figure S15E,F**). Vaccination was also associated with reduced cumulative viral replication in the nasal cavity (**Figure 3B**), and significantly lower peak viral loads (**Figure 3C**). However, only 50% of individuals with prior vaccination suppressed peak viral loads in the nasal cavity below 10^7^ copies/mL (**Figure S15G**), a presumably transmissible viral load.^24^ In the oral cavity, vaccination was not associated with earlier Type I or Type II IFN responses, nor significantly reduced viral loads (**Figure 3D-F**).

### Pre-infection COVID-19 vaccination was associated with nasal mucosal gene expression that *primed earlier Type II IFN responses and reduced viral replication*

To further interrogate the relationship between COVID-19 vaccination, Type II IFN response activation and viral replication in the nasal cavity, we evaluated a set of 12 genes canonically and uniquely associated with Type II IFN signaling and effector function (**Figure 4**). Among uninfected participants, 9 genes exhibited significantly higher basal expression in vaccinated than in unvaccinated individuals (**Figure 4A**), suggesting that IFN-γ-inducible genes are constitutively elevated by vaccination. Among infected participants, all 12 genes exhibited higher cumulative expression in the nasal cavities of vaccinated than unvaccinated participants during the 6 days preceding viral detection (**Figure 4B**). Longitudinal expression for each gene was aggregated among participants and stratified by vaccination status: genes responsible for pathway initiation such as *HLA-DRA, IFNG, IFNGR1* demonstrated higher basal expression for vaccinated than unvaccinated individuals, regardless of infection status, whereas genes responsible for cellular coordination and effector functions (*CXCL9, CXCL10, CXCL11, GBP1, GBP5, WARS1*) demonstrated earlier initiation following infection (**Figure 4C**). For all genes, cumulative early expression anti-correlated with virus shed by day 10 of infection, with *CXCL9* and *IFNGR1* exhibiting the strongest association (P_adj_<0.05), and strong relationships (P_adj_<0.1) observed for all other genes, except *HLA-DRA* (P_adj_=0.21) (**Figure 4D**). The relationship between higher early Type II IFN-related gene expression and lower viral replication was strongest when considering samples collected many days prior to detection of virus in the nasal cavity, with an effect that waned when considering samples collected just before viral detection (**Figure 4E**). These findings suggested that prior COVID-19 vaccination produced a transcriptional environment in the nasal mucosa prepared to initiate an early Type II IFN response and more effectively constrain viral replication.

**Figure 4.**
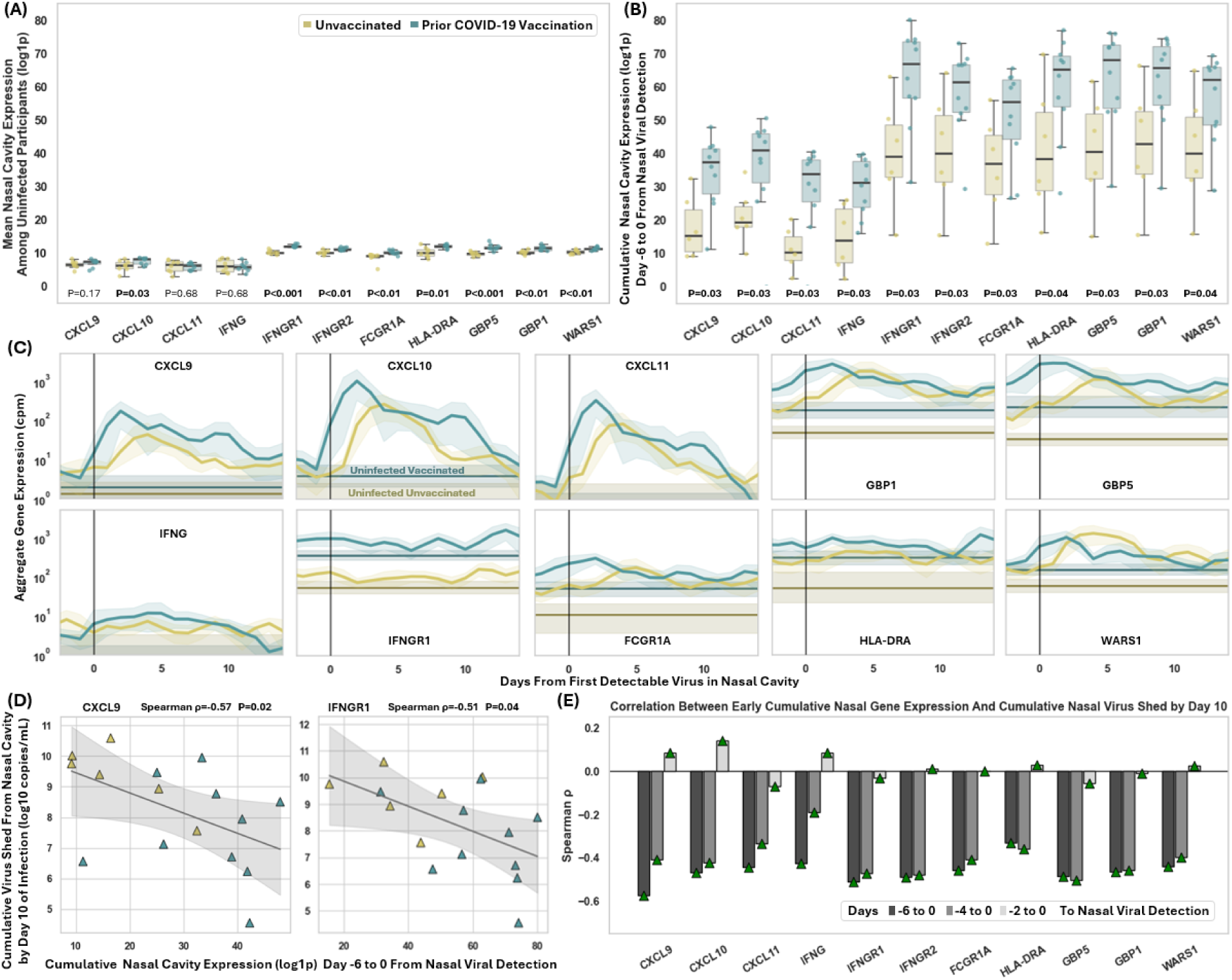
Pre-infection COVID-19 vaccination is associated with higher baseline expression and earlier upregulation of Type II IFN-related genes in the nasal cavity during early infection, which anticorrelates with viral replication in the nasal cavity. **(A)** For each uninfected participant, mean expression of each gene over time was calculated as a representation of basal expression, and the distribution of basal expression values was compared between participants who previously received a COVID-19 vaccine (blue) and unvaccinated participants (yellow). P values shown from Mann-Whitney U test with Benjamini Hochberg correction. **(B)** For infected participants, the AUC of expression measurements for each gene during the 6 days preceding viral detection (cumulative early expression) was calculated, and the distribution of cumulative expression was compared between vaccinated and unvaccinated participants. P values shown from Mann-Whitney U test with Benjamini Hochberg Correction). **(C)** For each day of infection, the mean expression of each gene was calculated for vaccinated or unvaccinated participants and plotted over time relative to detection of virus in the nasal cavity. Horizontal lines indicate mean basal expression among uninfected participants, either vaccinated (dark blue), or unvaccinated (dark yellow). Shading indicates 95% confidence intervals calculated via bootstrapping. **(D)** Cumulative early expression in the nasal cavity was plotted against cumulative virus shed from the nasal cavity by day 10 of infection. Unvaccinated participants are plotted as yellow triangles, while vaccinated participants are plotted as blue triangles, with Spearman correlation coefficient (ρ) and associated P value shown for each gene. Trendline was determined by Ordinary Least Squares, with shading indicating 95% confidence interval. **(E)** Similarly, Spearman correlation coefficients were calculated between cumulative early expression for varying time windows preceding detection of virus and cumulative virus shed. Anticorrelation indicates a relationship between greater early gene expression and lower cumulative viral replication.

### Tissue resident memory T cell signatures were higher among vaccinated participants, which *associated with earlier Type II IFN responses and reduced viral replication*

Tissue resident memory T cells (T_RM_) have been shown to initiate Type II IFN responses,^25,26^ but current COVID-19 vaccines have been shown to elicit only rare T_RM_ in the upper respiratory mucosa.^27–30^ We investigated the abundance of a T_RM_ signature in the upper respiratory mucosa among participants with or without prior COVID-19 vaccination, and whether the magnitude of T_RM_ signatures were related to the timing of mucosal Type II IFN responses and subsequent viral replication. To test this hypothesis, we calculated a T_RM_ module score for each sample based on a previously defined T_RM_ gene set^31^ and observed significant associations between prior COVID-19 vaccination, higher T_RM_ module scores, earlier initiation of Type II IFN responses, and reduced viral replication.

Vaccination was associated with higher T_RM_ module scores. Among nasal samples from uninfected individuals, vaccinated participants had significantly higher nasal cavity T_RM_ module scores than unvaccinated participants (**Figure 5A**), and in the first available nasal sample from all participants (regardless of subsequent infection), T_RM_ module scores were significantly higher among vaccinated individuals (**Figure 5B**). These associations were not observed in the oral cavities of uninfected participants (**Figure 5C**) or the first oral samples from all participants (**Figure 5D**). However, among infected participants, vaccinated individuals exhibited significantly higher T_RM_ module scores than unvaccinated participants during early infection periods in both the nasal and oral cavities (**Figure 5E,F**). Similarly, participants with any known SARS-CoV-2 antigenic exposure (either current infection or prior COVID-19 vaccination) exhibited significantly higher mean nasal cavity T_RM_ module scores than those without antigenic exposure (unvaccinated, uninfected participants) (**Figure S16A**).

**Figure 5.**
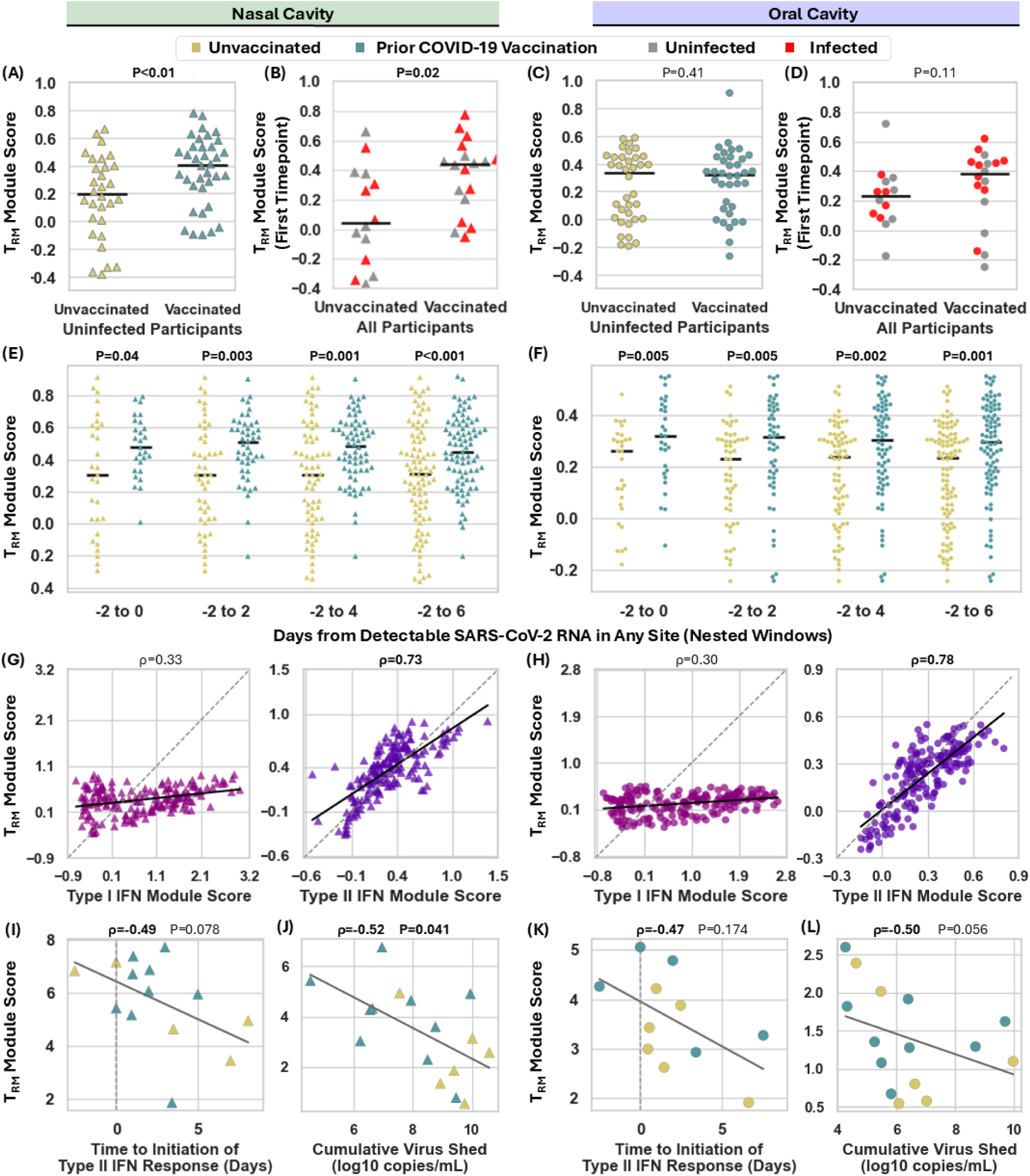
Pre-infection COVID-19 vaccination is associated with elevated nasal and oral T_RM_ module scores, earlier Type II IFN initiation, and reduced viral replication. T_RM_ module scores were calculated for each sample using a previously defined 112-gene tissue-resident memory T cell (T_RM_) signature.^31^ **(A,C)** Morning samples from uninfected participants were stratified by prior COVID-19 vaccination and compared by Mann Whitney U test with Benjamini-Hochberg correction. **(B,D)** First available samples from all participants were similarly stratified by vaccination status and compared. **(E,F)** Among infected participants, T_RM_ module scores across pre- and early-infection windows from days -2 to 6 were compared by vaccination status using Mann Whitney U test with Benjamini-Hochberg correction; horizontal lines indicate group means. **(G,H)** Nasal and oral T_RM_ module scores were compared with Type I and Type II IFN module scores by Spearman correlation. **(I,K)** Cumulative T_RM_ module scores from days -2 to 6 were plotted against time to Type II IFN initiation at each anatomical site. Type II IFN initiation was defined as the first site-specific timepoint exceeding an optimized infected-versus-uninfected threshold for the established Type II IFN gene set (GO:0034341,^21^ see **Figure S11**). **(J,L)** Site-specific cumulative TRM module scores from days -2 until first local viral detection were plotted against cumulative viral shedding through day 10. OLS regression is shown with Spearman’s ρ and *P*-value; color indicates vaccination status, but analyses were performed in aggregate.

Higher T_RM_ module scores were associated with earlier Type II IFN initiation and reduced viral replication. Despite very few genes overlapping between the T_RM_ and Type II IFN gene sets (**Figure S16B**), T_RM_ and Type II IFN module scores were strongly correlated in both the nasal and oral cavities (**Figure 5G,H**) and correlated significantly better than Type I IFN module scores (**Figure S16C**). Cumulative T_RM_ module scores during early infection were also strongly anticorrelated with the timing of Type II IFN response initiation (**Figure 5I,K**) and with cumulative virus shed by day 10 of infection (**Figure 5J,L**). These analyses suggest that COVID-19 vaccination elicited sufficient T_RM_ in the nasal mucosa to prompt earlier initiation of the local Type II IFN response and subsequently reduce viral replication.

## DISCUSSION

Upper respiratory mucosal surfaces are the primary sites of respiratory viral infection, replication and shedding, thus early immune responses that constrain viral replication at these sites can reduce the risk of onward transmission. Using human gene expression measurements from a unique set of paired samples from upper respiratory anatomical sites prospectively collected daily by individuals who became infected with SARS-CoV-2, we characterized the timing and spatial synchronization of early mucosal Type I and Type II IFN responses, their relative impacts on viral replication, and their modulation by prior COVID-19 vaccination.

Despite differences in early viral load trajectories among the nasal cavity, oral cavity and oropharynx of SARS-CoV-2 infected individuals, Type I IFN responses were initiated concurrently and often preceded detectable viral RNA in the nasal cavity. This result supports that testing for upregulation of nasal ISGs might enable earlier detection of viral infection to initiate earlier treatment or infection control measures.^32–35^ Among unvaccinated participants with sustained infection after SARS-CoV-2 challenge, ISGs were upregulated in the blood prior to the nasal cavity, suggesting a potential systemic route for synchronization among upper respiratory mucosal sites.^15^

Our analyses demonstrated that Type II IFN responses were not synchronized across upper respiratory mucosal sites within infected individuals, but earlier initiation was associated with lower local viral loads. Moreover, earlier initiation of Type II IFN responses was observed among participants with prior COVID-19 vaccination. Vaccinated individuals also exhibited subtle but constitutively elevated expression of IFN-γ-inducible and T_RM_ genes in the nasal cavity, which were further upregulated shortly after viral infection.

Our results fortify the link between enrichment of upper respiratory T_RM_s, earlier initiation of Type II IFN responses, and constraint of local viral replication in humans with naturally acquired SARS-CoV-2 infection. T_RM_s have a recognized role in limiting viral replication,^36–38^ including by rapidly producing IFN-γ upon SARS-CoV-2 exposure, as demonstrated in ex vivo nasal mucosal tissue^39^ and human lungs.^25^ In mice, airway CD4^+^ T cells recognizing SARS-CoV epitopes induced rapid local IFN-γ production and reduced disease burden.^40^ In a trial of non-human primates, IFN-γ induction by mucosal CD8^+^ T cells in the absence of SARS-CoV-2 neutralizing antibodies (NAbs) yielded similar reductions in nasal viral load as Nab-inducing vaccines.^41^ In human cross-sectional studies, higher magnitudes of a SARS-CoV-2-specific IFN-γ^+^ T cell response in venous blood were associated with reduced risk of infection^42^ and the presence of IFN-γ^+^ T cells in the upper airways was shown to prime viral recognition for enhanced viral clearance and local control of SARS-CoV-2 replication.^43^ Longitudinal measurements from a SARS-CoV-2 human challenge study of unvaccinated individuals further support the role of T cells in local viral clearance: CD4^+^ and CD8^+^ T cells infiltrated nasal mucosa within 1 day of inoculation for 9 participants with transient and abortive infections, but several days later among 7 participants with sustained infection.^15^ Establishment of T_RM_s in the upper respiratory mucosa and early activation of a Type II IFN response may constitute correlates of protection for vaccines intending to reduce viral replication and forward transmission.

Several studies have demonstrated that intramuscular COVID-19 vaccination can elicit or enhance early activation of T_RM_ populations in upper respiratory mucosal sites,^28,44–46^ though traditionally, intramuscular vaccines have not been very effective in establishing T_RM_ in the respiratory tract.^28–30,40,47–49^ Mucosal vaccines^50^ or hybrid strategies which follow intramuscular vaccination with an intranasal adjuvant (e.g. SARS-CoV-2 spike protein),^51–55^ have been shown to substantially augment the establishment of T_RM_s in the upper respiratory tract.^56^ Pre-existing cross-reactive T_RM_s may also contribute.^57^ Our study suggests that even low levels of T_RM_ responses generated by current COVID-19 vaccines can contribute to viral control, and the potential of vaccine strategies to augment establishment of mucosal T_RM_s, prompt early viral control, and block onward transmission.

Although earlier Type II IFN responses in both the nasal and oral cavities were associated with reduced viral replication, enrichment of T_RM_ signatures among vaccinated participants were more evident in the nasal than the oral mucosa, suggesting anatomic differences in current vaccine-induced mucosal immunity. We note that most studies assessing the establishment of T_RM_s in the upper respiratory tract have focused on the nasal cavity or nasopharynx^28,44,45^ and most mucosal vaccine candidates are administered only to the nasal cavity.^58^ Given that SARS-CoV-2 and other respiratory viruses can initiate infection at multiple upper respiratory anatomical sites, even complete protection of the nasal cavity may not prevent infection and transmission arising from the oral cavity or oropharynx. Our results suggest that future vaccine candidates intending to block breakthrough infection should have correlates of protection evaluated at multiple upper respiratory sites (e.g. nasal cavity, oral cavity, nasopharynx, oropharynx) where initial infection might occur. This study has several limitations that should be considered when interpreting our findings.

First, analyses were based on a modest number of participants with sustained incident infection (N=16), reflecting the logistical challenges of obtaining temporally dense, paired mucosal samples beginning before the onset of infection in humans. Comparable temporal resolution has largely been limited to controlled human challenge studies involving young, healthy, unvaccinated individuals inoculated with early SARS-CoV-2 strains.^15,33^ While our cohort included heterogeneity in age, vaccination status, health conditions, and viral variants, it was restricted to sustained infections and does not address transient or abortive infections. To our knowledge, however, daily mucosal immune profiling from multiple upper respiratory anatomical site starting before the onset of naturally acquired SARS-CoV-2 infection in humans has not previously been reported (**Table S2**).

Second, because vaccination occurred later in our study period, vaccinated participants were disproportionately infected with later SARS-CoV-2 variants, complicating efforts to disentangle vaccine effects from variant-specific biology. All participants providing oropharyngeal samples were vaccinated, precluding vaccine-related analyses for this site. Third, bulk RNA sequencing does not resolve cell-type-specific expression, nor directly quantify protein abundance or functional immune activity. While bulk mRNA sequencing captures tissue-level responses and cell populations underrepresented in single-cell datasets (e.g., neutrophils),^59^ we do not measure individual cells (including T_RM_s) or translated and secreted proteins, which are necessary to directly attribute IFN production and antiviral activity to specific mucosal cell populations. Functional profiling of upper respiratory resident T and B-cells would refine mucosal correlates of protection induced by vaccination.^60^ Finally, gene-set-based module scores may differentially reflect site-specific cellular composition, though our conclusions were robust across multiple independent Type I and Type II IFN gene sets.

In summary, our study highlights the key temporal association between local T_RM_, early induction of IFN-γ upon viral exposure, and delayed and reduced viral replication in the upper respiratory mucosa of vaccinated individuals.

## METHODS

### Design of a COVID-19 Household Transmission Study to Identify Individuals with Incident SARS-CoV-2 Infection, and Demographically Matched Uninfected Individuals

As previously described,^61^ between September 2020 and March 2022, individuals aged 6 and older residing in the Los Angeles area and sharing a household with a person recently diagnosed with SARS-CoV-2 were eligible to participate in our case-ascertained COVID-19 household transmission study (approved by the California Institute of Technology IRB, protocol #20-1026). Participants completed demographics and health questionnaires, as well as daily or twice-daily, paired, self-collected upper respiratory specimens (saliva and anterior nares swabs, and for participants enrolled after November 2021, also oropharyngeal swabs) collected in DNA/RNA Shield™ SafeCollect™ Swab Collection Kit (Zymo Research Inc., Catalogue #R1160-E) and Spectrum SDNA-1000 Collection Kit (Spectrum Solutions LLC, Draper, UT, USA). These upper respiratory specimens underwent testing for SARS-CoV-2 viral RNA using high analytical sensitivity RT-qPCR tests,^18,19^ with RT-ddPCR validation of viral load quantification methods as previously described.^18^

Among 415 participants from 116 households, we observed 161 (38.8%) individuals with laboratory-confirmed SARS-CoV-2 infection, 82 of whom were considered secondary cases within a household.^61^ Of these secondary cases, 16 participants were SARS-CoV-2-negative at enrollment but subsequently developed sustained infection; these individuals were classified as having naturally acquired incident SARS-CoV-2 infection, allowing longitudinal observation of infection dynamics across multiple upper respiratory anatomical sites from the earliest days of infection. From the 299 participants who did not develop SARS-CoV-2 infection, 16 uninfected control individuals were selected, matched on age, sex, COVID-19 vaccination status, and enrollment period (see **Supplemental Methods**). Using data and a total of 1,273 specimens from these 32 participants, we employed a case-control design to study longitudinal upper respiratory mucosal immune responses and viral load kinetics during acute SARS-CoV-2 infection.

### Generation and Pre-Processing of Human Gene Expression Data

In addition to longitudinal viral load quantification (previously described^18,19^), longitudinal human gene expression was measured using an optimized and validated human mRNA sequencing pipeline (described further in **Supplemental Methods**). Briefly, RNA was isolated using the Quick-DNA/RNA™ HT kit (Zymo Research Corporation, Catalogue Number R2150) with a DNase I digestion step to remove DNA. RNA was converted into cDNA using the lllumina RNA Prep with Enrichment (L) Tagmentation kit (Illumina, Inc., Catalogue Number 20040537) and cDNA libraries were subsequently enriched for human mRNA transcript sequences using the Twist Bioscience for Illumina Exome 2.5 Panel. Resulting human exome-enriched cDNA libraries were pooled and sequenced to a target depth of 25 million reads on a NovaSeq 6000 platform. Samples which did not initially achieve quality control standards for RNA extraction and DNA digestion, cDNA library generation, human sequence enrichment or sequencing depth underwent modified protocols, described in **Supplemental Methods**. Samples not meeting quality control standards following modified protocols were excluded from analysis.

Raw RNA sequencing data was mapped to human GRCh38.p13^62^ reference using STAR v2.7.11b^63^ with default parameters, and pre- and post-mapping quality control assessment by MultiQC v1.27.1^64^ and Qualimap 2 v2.3^65^ respectively. Raw gene counts were extracted using featureCounts v2.22.1.^66^ Samples with fewer than 10 million human-mapping raw gene counts were excluded from analysis due to insufficient coverage. Raw gene counts were then normalized by specimen type group using the trimmed mean of M-values with singleton pairing^67^ implemented in edgeR.^68–70^ Normalized gene expression values were merged with sample metadata for subsequent analyses.

### Statistical Analyses

For longitudinal analyses, time was represented either as days relative to study enrollment (for uninfected participants) or to the first detectable SARS-CoV-2 RNA in any anatomical site within each infected participant. Longitudinal trajectories (viral load, individual genes, or module scores) were summarized within stated time windows by pooling available sample-level observations after rounding time to the nearest day, with 95% confidence intervals estimated for each window by percentile bootstrap with 1000 resamples. Samples with no detectable virus (ND) were imputed to a value of 1 copies/mL, and samples with detectable but not quantifiable viral loads (NQ) were imputed to a value of 10 copies/mL. Cumulative viral load was calculated using trapezoidal integration of linear-scale viral load measurements from a given participant and anatomical site, then log10-transformed for downstream analyses. Trapezoidal integration was also applied to linear-scale, normalized gene expression measurements to calculate cumulative gene expression within given infection windows, then log1p-transformed for downstream analyses. Module scores for pre-defined gene sets representing relevant immune programs (Type I, II, and III IFN, and tissue-resident memory T cells) were computed from the log1p-transformed normalized bulk RNA-seq expression matrix using a control-gene-subtracted approach: for each sample, the mean expression of genes in the target set was calculated and compared with the mean expression of a pooled expression-matched control background. For each target gene, up to 1,000 control genes were randomly sampled without replacement from genes with mean expression within 90-110% of the mean expression of the target gene. The mean expression of the pooled sampled control genes was then subtracted from the mean expression of the target gene set to yield the module score. Where stated, module scores were min-max normalized within participant time courses prior to visualization. For each gene set and anatomical site, the initiation threshold was defined as the lowest module score achieving >90% sensitivity and >75% specificity for distinguishing infected from uninfected participants. Initiation was defined as the first timepoint at which a participant’s module score exceeded this threshold.

For temporal correlations between viral load, gene expression, or module scores within a participant, the Pearson correlation was calculated among exact matched timepoints, otherwise Spearman rank correlation coefficients were calculated between continuous variables with either linear regression or ordinary least-squares trend lines added for visualization. In some analyses, correlation coefficients were compared to coefficients generated by a null model with randomly permuted time or sample labels using Wilcoxon signed-rank tests.

Differences in cumulative viral load, peak viral load, timing differences, and other participant-level summaries between groups were tested using Mann-Whitney U tests. Paired comparisons, such as local-virus versus first-virus timing intervals within the same participants, were tested using Wilcoxon signed-rank tests. P-values were adjusted for multiple comparisons using either Bonferroni or Benjamini-Hochberg within each figure or analysis family. Statistical testing was implemented in Python 3.12.4 using SciPy v1.14.1.

## Data Availability

All data supporting the findings of this study are available as described below. Underlying data presented in this manuscript and de-identified, processed bulk RNA-seq data with associated metadata are available at https://doi.org/10.22002/fh69y-nef75. Code used for data processing and analysis is available at https://github.com/IsmagilovLab/COVIDHostAnalysis. Raw RNA sequencing data are available upon reasonable requests and in accordance with pertinent restrictions related to human subject privacy and informed consent.

## ACKNOWLEDGEMENTS

The authors wish to thank Dr. Natasha Shelby, Dr. Karen Heichmann, Dr. Valerie Arboleda, Dr. Robert Schlaberg, and Dr. Dan Wattendorf for suggestions and feedback on study design and specimen processing methods. We wish to thank Dr. Andy Lim, Dr. Reid Akana, and Dr. Greg Fedewa for suggestions related to biostatistical considerations and approaches to analysis of this dataset. We wish to thank Dr. Mildred Galvez, Dr. Prashant Bhat, Dr. Delaney Sullivan, Dr. Laura Leubbert, Dr. Lior Pachter and Hengyu Wang for feedback and recommendations regarding quality assessment and processing of RNA sequencing data, and provision of HeLa cell culture lysate from Dr. Prashant Bhat. We wish to thank Dr. Michael K. Porter and Dr. Minkyo Lee for expertise when validating methods for quantification of human mRNA targets by RT-qPCR. We also wish to thank Dr. Ellen Rothenberg and Dr. Leonard Warren for suggestions to best interpret the biological and immunological signals within this dataset. We also wish to thank We also wish to thank Chih-Yu Liu, Michael Adrian Rubio, Jennifer Telish, Sophia Gonzales, and Danny Thai at Zymo Research Corp. for carefully and rigorously implementing specimen processing protocols. We thank Mingda Jin and Suresh Gupta for assistance transferring, securing and storing raw sequencing data files for analysis.

