## Supplemental Information for "Early Mucosal Type II Interferon Limits SARS-CoV-2 Replication in Humans"

### Contents


1. **Supplemental Methods**
    1. Viral Load Quantification
    2. Participant Demographics and Classifiers
    3. Case-Control Matching Algorithm and Specimen Selection
    4. Additional Method Details of Gene Expression Data Generation
    5. Additional Details of Statistical Analyses
2. **Supplemental Data**
    1. Table S1
    2. Figure S1
    3. Figure S2
    4. Figure S3
    5. Figure S4
    6. Figure S5
    7. Figure S6
    8. Figure S7
    9. Figure S8
    10. Figure S9
    11. Figure S10
    12. Figure S11
    13. Figure S12
    14. Figure S13
    15. Figure S14
    16. Figure S15
    17. Figure S16
    18. Table S2
3. **Author Contribution Statements and Contact Information**

### Supplemental Methods

*Viral Load Quantification*

SARS-CoV-2 viral loads were quantified using FDA-authorized RT-qPCR assays which we independently validated to have Limits of Detection at or below 1000 copies/mL.[1,2] To convert from SARS-CoV-2 *N* gene Ct values to viral loads in genomic copy equivalents (copies) per mL of specimen, we generated a 13-point standard curve (dynamic range 250 to $1x10^9$ copies/mL) of contrived specimens created from sample matrix from SARS-CoV-2 negative participants and heat-inactivated viral particles (BEI Cat. N4-52286 Lot 70034991) or a high viral load specimen previously quantified by both RT-qPCR and RT-ddPCR. Positive specimens with viral loads that would be quantified below the assay LOD were considered detected but not quantifiable (NQ). Specimens with no detected SARS-CoV-2 target amplification or Ct values >40 were considered not detected (ND).

*Participant demographics and classifiers*

All participants completed a health questionnaire at study enrollment. Participant sex was classified based on self-report in response to the question, "What sex were you assigned at birth?" with response options including Male, Female, or Other (free-text). Reported age at enrollment was converted into the following predefined age categories: 6-12, 13-20, 21-30, 31-40, 41-50, 51-60, 61-70, and ≥71 years. Participants reported medical history through structured questions targeting conditions of relevance to the study (e.g., asthma), supplemented by free-text fields allowing reporting of additional conditions. Medication and supplement use was assessed using indication-based categories (e.g., allergy medications, immunosuppressive medications, psychiatric medications, inhaled medications), followed by free-text entry of specific agents and an open-response field to list all medications. Attention-check items were embedded within these sections to ensure participant engagement. Free-text responses describing medical conditions and medications were reviewed and systematically coded by a research team member trained in medical terminology to enable standardized inter-participant comparisons. Participants who reported receipt of at least one dose of a COVID-19 vaccine authorized for use by the U.S. Food and Drug Administration >14 days prior to enrollment were classified as vaccinated; participants reporting no prior COVID-19 vaccination were classified as unvaccinated.

*Case-Control Matching Algorithm and Specimen Selection*

Each participant with incident SARS-CoV-2 infection (Case) was paired with a matched uninfected participant (Control). For each infected participant, the uninfected participant cohort was strictly subset by matching sex and age range (defined above). Vaccination histories were reviewed, and participants who had received at least one dose of a COVID-19 vaccine authorized by the U.S. FDA at least 14 days prior to enrollment were classified as vaccinated. The uninfected participant cohort was then strictly subset to matching vaccination status. Potential matches were further subset to participants for whom at least 5 samples from each anatomical site spanning 5 day of collection were available. For most participants, only one potential match was available. When multiple potential matches were available, manual review of smoking history, active medications, medical comorbidities, and self-reported health status were used to downselect to an optimal match. Samples from Cases and Controls underwent processing for human gene expression measurements as described below.

*Additional Method Details of Gene Expression Data Generation*

Selected specimens were thawed from -80°C to room temperature, and 300µL of primary specimen volume was aliquoted into designated KingFisher 96-deep-well plates compatible with automated processing on a KingFisher Flex (Thermo Fisher Scientific, Catalogue #95040450).

Each plate contained specimens of a given specimen type (e.g. all nasal swab, or all saliva, or all oropharyngeal swab) from the same study phase (e.g. all Phase I specimens, or all Phase II specimens). To minimize the potential for longitudinal measurements from a given participant and anatomical site to be confounded by processing batch effects, specimens from a given participant and specimen type were assigned to be processed within a single plate. Prior to aliquoting specimen volume to pre-defined wells on each processing plate, a primary operator arranged primary specimen types per the plate map, and a second operator independently confirmed the arrangement of the specimens. As a third-pass check, the primary operator re-confirmed specimen placement prior to aliquoting each specimen. In addition to primary specimen volumes, every plate contained 300μL of nuclease free water as a negative control, 300μL volume from a single-stock of HeLa cell lysate in DNA/RNA Shield as a positive control, and 300μL volume from a single-stock of saliva in DNA/RNA Shield pooled from 10 unique participants (not within the SII or TN cohorts) as a positive control in a relevant, challenging clinical specimen matrix.

Target specimens pre-aliquoted into 96-well plates underwent automated RNA extraction and purification via the Quick-DNA/RNA HT kit (Zymo Research Corp, Catalogue #R2150), including DNase I digestion, on a KingFisher Flex Purification System (Thermo Fisher Scientific). RNA was eluted in 50μL of nuclease-free water. 2μL of eluate was quantified using the Qubit 1X dsDNA HS Assay to ensure successful DNA digestion (Invitrogen, Catalogue #Q33231). Prior to processing target specimens, technical control specimens were processed in this automated platform in triplicate and checkerboard pattern to confirm technical reproducibility and absence of well-to-well cross contamination. For target specimens, 5μL of elution volume underwent RT-qPCR (TaqMan™ Fast Virus 1-Step Master Mix for qPCR, ThermoFisher Scientific Catalogue #4444434) for two human mRNA-specific targets in the MYH9 and ACTB genes (ThermoFisher

Scientific, Hs01066381_m1 with FAM fluorophore, and ThermoFisher Scientific Hs99999903_m1 with VIC fluorophore). For specimens with Ct values below 35 for both human mRNA targets, 8.5μL of elution volume underwent library preparation (Illumina RNA Prep with Enrichment (L) Tagmentation, Illumina Catalogue #20040537) with 17 amplification cycles. cDNA libraries were quantified by Qubit 1X BR DNA (Invitrogen, Catalogue #Q33236); select, representative samples underwent library fragment size distribution quantification (Agilent D1000 ScreenTape on Agilent TapeStation 2200, Catalogue #50675582). Triplex pools for human exome enrichment were created by pooling 200ng of cDNA from each constituent libraries, originating from specimens collected by the same participant when possible. 7.5μL of select cDNA libraries with concentrations incompatible with triplex pooling (ie. <30ng/μL) were underwent singleplex enrichment. Enrichment (Illumina RNA Prep with Enrichment (L) Tagmentation, Illumina Catalogue #20040537) was performed using the Twist Exome 2.5 Plus panel (Illumina Catalogue #20076914) and enriched libraries amplified for 14 cycles. Enriched libraries were pooled, and sequenced paired-end 2x150bp on the NovaSeq 6000, targeting 40 million reads per sample for oral cavity specimens, and 25 million reads per sample for nasal cavity and oropharyngeal specimens.

A subset of specimens did not achieve quality control metrics during initial processing. Specimens which did not achieve human MYH9 Ct value below 40 following RNA extraction were excluded from subsequent processing. Specimens which did not achieve a minimum of 30ng/μL of cDNA following an initial library preparation attempt were re-attempted, with 21 amplification cycles. Human exome enrichment pooling (singleplex, duplex, or triplex) was then tailored to the available cDNA concentration for these samples. For samples which did not achieve target number of human reads on initial sequencing, either the human exome enriched pool containing the sample was re-sequenced, or re-enrichment was performed prior to re-sequencing. FASTQ reads from samples which underwent repeat sequencing attempts were concatenated upstream to sequencing data quality control steps, human genome mapping and gene expression quantification. Samples which failed to achieve a minimum of 10 million mapped human reads after this modified protocol were excluded from subsequent analysis, to prevent variation driven by technical factors (sequencing depth) versus biological effects.

*Additional Details of Statistical Analyses*

Unless comparing anatomical sites, differential gene expression analyses were performed separately by anatomical site, using DESeq2 implemented in R v4.4.1 applied to raw gene count data with default size-factor normalization and dispersion estimation. Gene-wise counts were modeled with a negative binomial generalized linear model using the categorical variable of interest (e.g. Sex) specified as the design factor, with log2-fold change estimated by the model, and p-values from Wald tests adjusted for multiple hypothesis testing by Bonferroni correction.

Principal Component Analysis (PCA) was performed on TMM normalized abundance values of protein-coding genes in human upper respiratory specimens (nasal cavity, oral cavity, oropharynx). Samples failing quality control were excluded. Genes with zero abundances across all retained samples were removed, as were genes in the lowest 10% of mean abundance among all samples. To reduce noise and identify informative genes, the relationship between log-transformed coefficient of variation and log-transformed mean abundance for each gene was modeled using locally weighted scatterplot smoothing (LOWESS; smoothing fraction=0.3), and residual variability was calculated as the difference between the observed log coefficient of variation and the LOWESS-fitted value. Genes were retained if their residual variability was at least 2.5 standard deviations above the mean residual variability. Among remaining genes, a pseudocount of 0.5 was added before natural logarithm transformation. Each gene (feature) was then standardized to zero mean and unit variance using StandardScaler from scikit-learn, and PCA was performed using sklearn.decomposition.PCA with 10 principal components. Sample scores were obtained from the fitted PCA model and used for visualization.

Single sample Gene Set Enrichment Analysis[3] was performed on log1p-transformed, normalized gene expression data, using ssGSEAProjection (v4) on GenePattern[4] using KEGG Medicus database pathways and default parameters for weighting exponent, minimum gene set size, and combine mode.

To estimate the relative abundance of cell types in each sample, we created a signature matrix using publicly available scRNA-seq data generated from nasal swab specimens from participants with SARS-CoV-2 infection[5] and the FindMarkers function from Seurat[6,7] run in R v4.4.1. Using this signature matrix, cell type abundances were deconvoluted using CIBERSORTx[8] with default parameters and linear-scale, normalized gene expression data as the mixture file.

Time-to-event analyses comparing vaccination groups or sex groups for IFN initiation relative to viral-load threshold crossing were performed using Kaplan-Meier curves and log-rank tests.

### Supplemental Data

(TABLE ON NEXT PAGE) **Table S1. Individual-Level Participant Attributes.** Cohort of Participants from the Caltech COVID-19 Study for whom specimens underwent human mRNA sequencing. Participants with Participant ID (PID) values that begin with "P" were enrolled in Phase I while those that begin with "Z" were enrolled in Phase II of the study. "Case" indicates that the participant had RT-qPCR diagnosed sustained incident SARS-CoV-2 infection, while the subsequent demographically-matched "Control" was negative for SARS-CoV-2 RNA by a high analytical sensitivity RT-qPCR test (see **Methods**). "Sex" reports response to sex assigned at birth, with "M" for Male, "F" for Female. No participants reported Other (free-text). Participants also reported gender identity (Man/Boy, Girl/Woman); all participants reported cis-gender identity to sex at birth, with no participant reporting Other (free-text). "COVID-19 Vax" was determined by participant self-report, and classified as either "Unvax", or "Vax" if they reported at least one dose of an authorized COVID-19 vaccine primary series ("Moderna" authorized as "Spikevax" with prior investigations under the name "mRNA-1273", or "Pfizer" being the "Pfizer-BioNTech mRNA COVID-19 vaccine" authorized under the name "Comirnaty" and prior investigations under the name "BNT162b2", and "Janssen" or "J&J" being the "Johnson & Johnson COVID-19 vaccine" authorized under the name "Jcovden" and prior investigations under the name "Ad26.COV2.S". All "Unvax" participants were infected with viral variants ancestral to the SARS-CoV-2 Delta variant, and all "Vax" participants were infected with the SARS-CoV-2 Omicron variant, determined by SARS-CoV-2 viral sequencing.[9] "Oral", "Nasal" and "OP" (Oropharyngeal) specimens refer to the number of specimens of each type for which mRNA sequencing was attempted. "Active Meds" list medications reported as actively taking by the participant; generic drug names are listed where possible, when either brand or generic name was provided by the participant, otherwise drug category. Due to heterogeneity in reported names, "Vitamin" was condensed to a single drug category, though most participants reported a form of daily multivitamin or Vitamin C supplement. "Medical History" provides comorbidities listed by the patient; "T2DM" is Type 2 Diabetes Mellitus, "HTN" is hypertension. "Smoking history" reports tobacco smoking or vaping of any substance; "Never" is listed if the participant reported never smoking tobacco or vaping; "Former" is if the patient reported previous tobacco smoking or vaping. "Health State" is a self-reported 5-point Likert scale of baseline health status with options Poor (1), Fair (2), Good (3), Very Good (4), and Excellent (5). "Fig S1 Panel" refers to the data shown in each panel of **Figure S1**, as well as corresponding panels in **Figure S7** and **Figure S12**. "Match" refers to the Case or Control participant age-, sex-, and COVID-19-vaccination status-matched to the given participant.

(TABLE CAPTION ON PRECEDING PAGE)

| ID | GROUP | AGE BIN | BIRTH SEX | COVID-19 VAX | ORAL (N) | NASAL (N) | OP (N) | ACTIVE MEDS | MEDICAL HISTORY | SMOKING HISTORY | HEALTH STATE | FIG S1 PANEL | MATCH |
|---|---|---|---|---|---|---|---|---|---|---|---|---|---|
| **PA** | Case | [50-60) | M | Unvax | 27 | 28 | 0 | Fluticasone propionate, Acetaminophen | Obesity | Former (Current Vape) | 3 | **A** | **PQ** |
| **PB** | Case | [0-10) | F | Unvax | 31 | 36 | 0 | Vitamin | None | Never | 5 | **B** | **PR** |
| **PC** | Case | [50-60) | M | Unvax | 37 | 38 | 0 | Vitamin | None | Former | 2 | **C** | **PS** |
| **PD** | Case | [30-40) | M | Unvax | 29 | 28 | 0 | None | T2DM | Never | 2 | **D** | **PT** |
| **PE** | Case | [50-60) | F | Unvax | 38 | 35 | 0 | Acetylsalicylic acid, Vitamin | None | Never | 4 | **E** | **PU** |
| **PF** | Case | [30-40) | F | Unvax | 36 | 38 | 0 | Oral contraceptive, Vitamin | None | Never | 3 | **F** | **PV** |
| **PG** | Case | [10-20) | F | Unvax | 43 | 42 | 0 | Vitamin | None | Never | 3 | **G** | **PW** |
| **ZH** | Case | [40-50) | M | Vax | 21 | 21 | 22 | None | None | Never | 5 | **H** | **ZX** |
| **ZI** | Case | [0-10) | M | Vax | 28 | 27 | 27 | None | None | Never | 5 | **I** | **ZY** |
| **ZJ** | Case | [0-10) | M | Vax | 14 | 14 | 14 | Acetaminophen | None | Never | 5 | **J** | **ZZ** |
| **ZK** | Case | [30-40) | M | Vax | 27 | 27 | 27 | None | None | Former | 4 | **K** | **ZAA** |
| **ZL** | Case | [20-30) | M | Vax | 8 | 7 | 8 | Antibiotic ear drops (Unspecified) | None | Never | 2 | **L** | **ZAB** |
| **ZM** | Case | [20-30) | F | Vax | 11 | 11 | 11 | Oral contraceptive, Vitamin | None | Never | 3 | **M** | **ZAC** |
| **ZN** | Case | [30-40) | F | Vax | 22 | 22 | 22 | Vitamin | Anxiety | Never | 3 | **N** | **ZAD** |
| **ZO** | Case | [40-50) | F | Vax | 21 | 21 | 21 | Vitamin | None | Never | 5 | **O** | **ZAE** |
| **ZP** | Case | [30-40) | F | Vax | 18 | 18 | 18 | Allergy | None | Never | 2 | **P** | **ZAF** |
| **PQ** | Control | [50-60) | M | Unvax | 11 | 11 | 0 | Vitamin | None | Never | 4 | **Q** | **PA** |
| **PR** | Control | [0-10) | F | Unvax | 13 | 13 | 0 | Vitamin | None | Never | 4 | **R** | **PB** |
| **PS** | Control | [50-60) | M | Unvax | 10 | 11 | 0 | Vitamin | None | Never | 4 | **S** | **PC** |
| **PT** | Control | [30-40) | M | Unvax | 6 | 7 | 0 | Vitamin | None | Former | 4 | **T** | **PD** |
| **PU** | Control | [50-60) | F | Unvax | 11 | 11 | 0 | Antihypertensive (Unspecified), Vitamin | HTN | Never | 2 | **U** | **PE** |
| **PV** | Control | [30-40) | F | Unvax | 12 | 14 | 0 | Vitamin | None | Never | 4 | **V** | **PF** |
| **PW** | Control | [10-20) | F | Unvax | 10 | 10 | 0 | None | None | Never | 5 | **W** | **PG** |
| **ZX** | Control | [40-50) | M | Vax | 8 | 8 | 9 | Vitamin | None | Former | 4 | **X** | **ZH** |
| **ZY** | Control | [0-10) | M | Vax | 8 | 8 | 8 | None | None | Never | 5 | **Y** | **ZI** |
| **ZZ** | Control | [0-10) | M | Vax | 8 | 10 | 8 | None | None | Never | 3 | **Z** | **ZJ** |
| **ZAA** | Control | [30-40) | M | Vax | 9 | 9 | 9 | None | None | Never | 5 | **AA** | **ZK** |
| **ZAB** | Control | [20-30) | M | Vax | 5 | 5 | 5 | None | None | Never | 2 | **AB** | **ZL** |
| **ZAC** | Control | [20-30) | F | Vax | 7 | 6 | 6 | Acetaminophen, Ibuprofen | None | Never | 3 | **AC** | **ZM** |
| **ZAD** | Control | [30-40) | F | Vax | 6 | 7 | 6 | None | None | Former | 3 | **AD** | **ZN** |
| **ZAE** | Control | [40-50) | F | Vax | 11 | 11 | 10 | None | None | Never | 4 | **AE** | **ZO** |
| **ZAF** | Control | [30-40) | F | Vax | 6 | 6 | 6 | None | None | Never | 2 | **AF** | **ZP** |

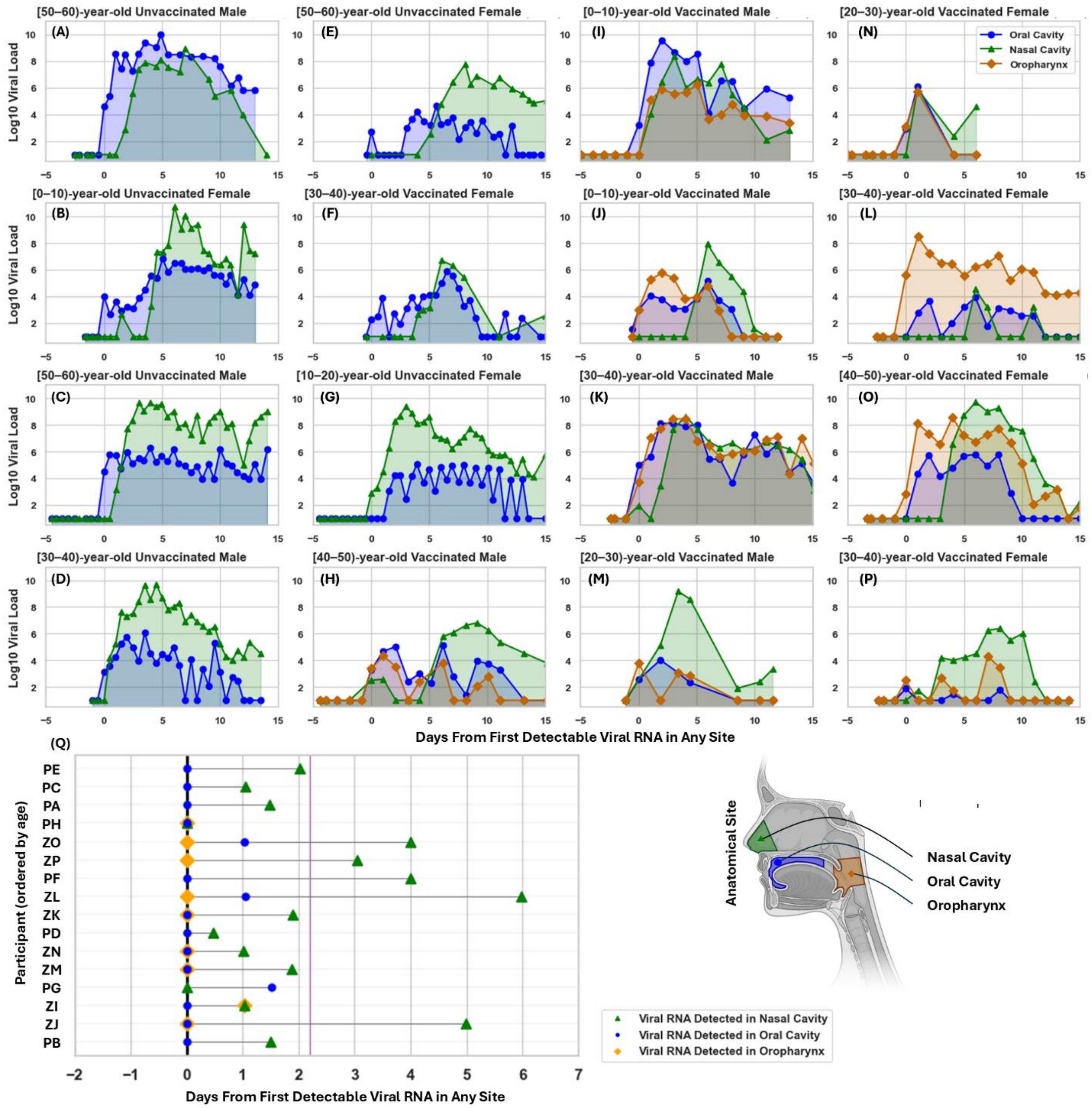


**Figure S1. Longitudinal, quantitative viral load measurements from each anatomical site from each individual over time**. Viral load data by individual participant are shown in each panel **(A-P)** relative to time from first positive result in any anatomical site within that participant. Participant age range and sex is shown above each panel. Data shown in these plots has been previously reported.[1,2] **(Q)** For each participant, the order of positivity among anatomical sites is shown on a timeline. Purple vertical line shows the average delay from first detectable viral RNA in the oral cavity or oropharynx to first detectable viral RNA in the nasal cavity.

***High quality human mRNA sequencing from daily self-collected specimens from the oral cavity, nasal cavity, and oropharynx enable robust quantification of mucosal gene expression***

Self-collected samples exhibited only minor sampling variation over time (**Figure S2A**). Human MYH9 Ct values were highly stable within samples from an individual over time, with a standard deviation of longitudinal MYH9 Ct values less than 3 Ct for 100% of participants collecting saliva and throat swab specimens, and 90.3% of participants collecting nasal swabs (**Figure S2B-C**). Despite physical sample complexity and concerns of RNA quality, cDNA libraries were successfully generated from self-collected upper respiratory human clinical specimens (**Figure S2D**). 100.0% (556 of 556) of oral cavity specimens, 97.9% (474 of 484) nasal cavity specimens, and 99.6% (232 of 233) oropharyngeal specimens achieved cDNA library concentrations sufficient for sequencing. The method successfully enriched for human transcripts, and yielded greater than 10 million human reads per sample in 96.2% of oral, 84.7% of nasal, and 94.4% of oropharyngeal samples (**Figure S2E**).

The sample processing pipeline generated robust measurements of gene expression. Gene expression was highly correlated (Pearson $r > 0.92$) between replicates of select human clinical samples ($n = 3$) that underwent duplicate extraction, library preparation, human exome enrichment, sequencing and data processing (**Figure S2F**). Additionally, each of 17 processing batches included a technical replicate of HeLa cell lysate and of pooled volume from oral cavity specimens from healthy human donors. Expression of select housekeeping genes was highly consistent among the replicates of each technical control sample type between preparation batches (**Figure S2G**). For genes with low abundance (e.g. mean normalized expression < 0.01), coefficients of variation increased, consistent with stochastic detection. Among genes with mean normalized expression above 0.01 (>93% of all genes), 85% or more had coefficients of variation less than 1 (**Figure S2H**).

Gene expression measurements exhibited anticipated biological patterns. Genes with expected differences in expression based on biological sex (e.g. *UTY, ZFY, EIF1AY, RPS4Y1, KDM5D, USP9Y, TSPY9*) indeed exhibited significant differential expression based on participant-reported sex, for each anatomical sampling site (**Figure S2I**). Relatedly, samples clustered strongly by anatomical site, consistent with expected tissue-dependent differences in gene expression (**Figure S2J**)

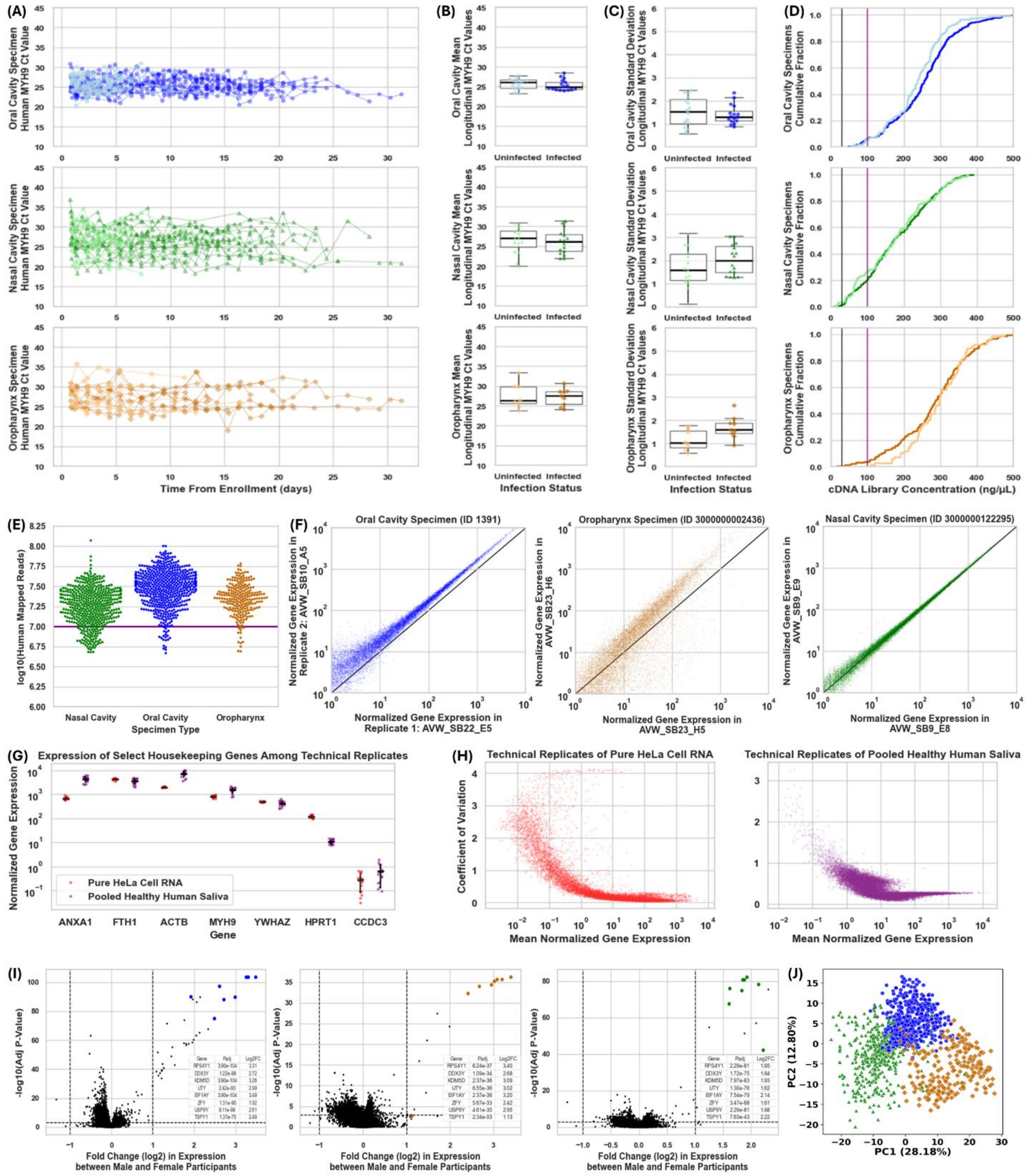


(FIGURE CAPTION ON SUBSEQUENT PAGE)

(FIGURE ON PRECEDING PAGE) **Figure S2. Daily self-collected oral, nasal and oropharyngeal specimens successfully generated high quality human mRNA sequencing and gene expression data. (A)** Human MYH9 expression measured by RT-qPCR following RNA extraction from each sample. Lines demonstrate raw MYH9 Ct values from each sample collected by a given participant either without (lighter color) or with (darker color) acute SARS-CoV-2 infection, for each anatomical site sampled (oral cavity, blue; nasal cavity, green; oropharynx, orange). **(B-C)** Box and whisker plots show the distribution, while each datapoint represents the mean **(B)** or standard deviation **(C)** of MYH9 Ct values for specimens collected by a given participant over time, for those without (lighter color) and with (darker color) SARS-CoV-2 infection, for each anatomical site sampled (oral cavity, blue; nasal cavity, green; oropharynx, orange). **(D)** Following RNA extraction with DNA digestion, cDNA libraries were generated and quantified. The empirical cumulative distribution of cDNA library concentrations for participant without (lighter color) or with (darker color) acute SARS-CoV-2 infection are shown for each anatomical site sampled (oral cavity, blue; nasal cavity, green; oropharynx, orange). Vertical black line indicates the minimum cDNA library concentration required for the subsequent step of human exome enrichment (30ng/μL), and the vertical purple line indicates a robust cDNA library concentration (100ng/μL). **(E)** Enriched cDNA libraries generated from each sample were pooled and sequenced. Resulting sequencing data underwent mapping to the human reference genome to yield the total raw count of human mapped reads for each sample. The horizontal purple line indicates sufficient depth for robust gene expression analysis (10 million raw human reads per sample). **(F)** A subset of primary specimens from each anatomical site underwent preparation in duplicate. Normalized expression for each gene in each replicate were plotted against each other. Black line indicates identity. **(G)** Volume from pooled HeLa cell culture lysate (red) and pooled oral cavity specimen volume from 10 healthy participants (purple) were included as consistent technical controls within each sample processing batch. HeLa cell lysate was included as a pure, high quality RNA control, whereas pooled oral cavity specimen volume which is expected to have a complex microbial background and lower quality human mRNA RNA was included to approximate other self-collected upper respiratory human clinical specimens. Normalized expression values for select housekeeping genes with varying levels of expression are shown for each replicate, of each control sample type. Horizontal black bars indicate the median normalized gene expression among replicates of that control sample type. **(H)** For each gene, the coefficient of variation is plotted against the mean normalized gene expression among technical replicates of each control sample type. **(I)** Samples from each anatomical site were stratified by participant-reported sex assigned at birth, and differential expression analysis was performed. Each datapoint represents the log2 fold change between male and female participants versus the -log10 of Bonferroni corrected P-value. A priori defined genes expected to exhibit sex-dependent differences in gene expression are colored by anatomical site on each plot. **(J)** Principal Component Analysis was performed on normalized gene expression data from all human clinical samples, with the first two principal components plotted. Datapoints are colored by anatomical site. Axis labels reported the percentage of total variance explained by PC1 and PC2, respectively, as calculated from the PCA explained variance ratios.

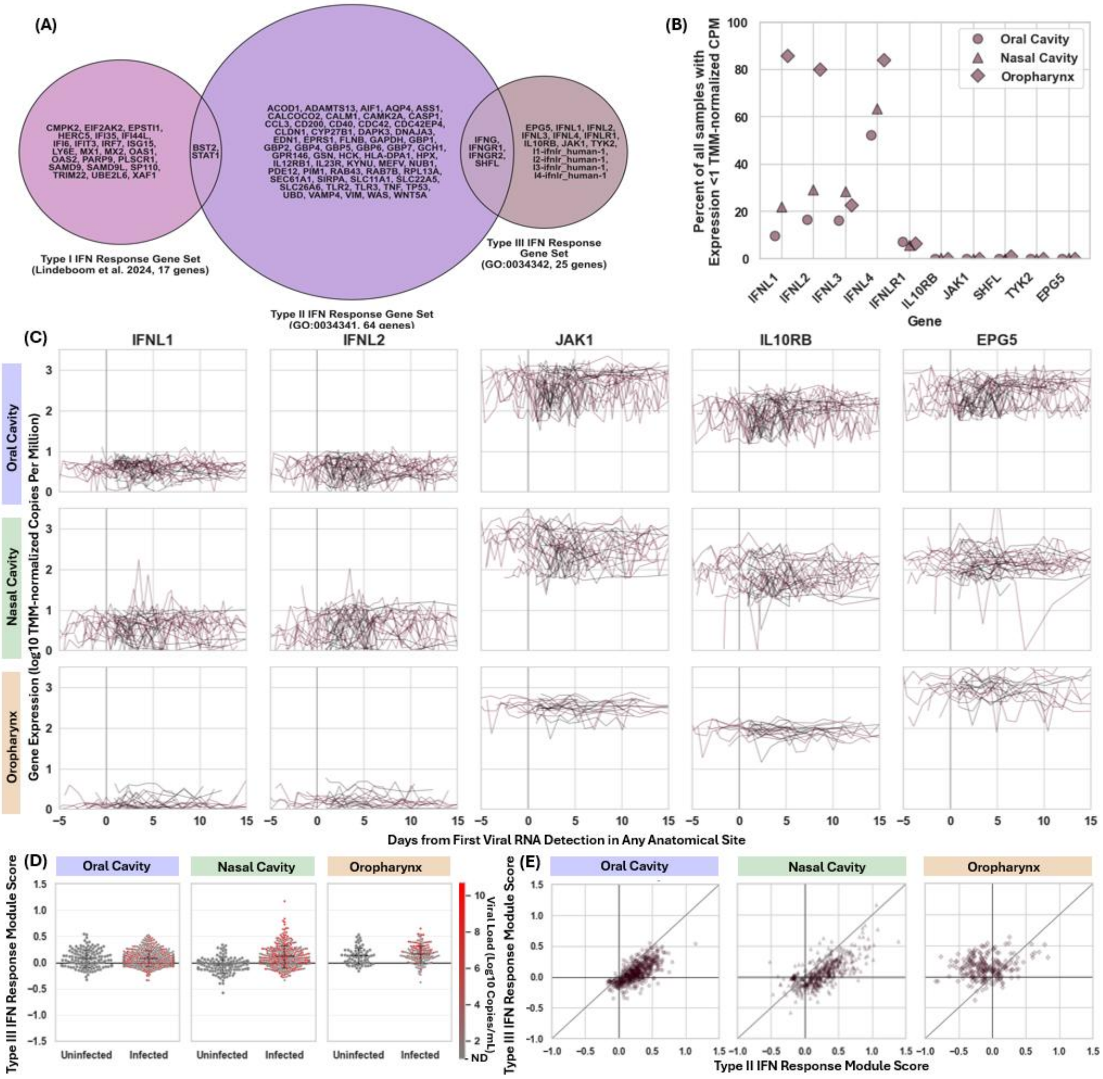


**Figure S3. The Type III IFN Response Gene Set is non-specific, lowly expressed, and not differentiable between infected and uninfected participants.** For each sample, a module Score was calculated using the 19 genes present in the "response to type III interferon" pathway from AmiGO (GO:0034342[10]). **(A)** Genes included in the Type III IFN response include *IFNL1, IFNL2, IFNL3, IFNL4, IFNLR1, IL10RB, JAK1, SHFL, TYK2, EPG5, IFNG, IFNGR1, IFNGR2, l1-ifnlr_human-1, l2-ifnlr_human-1, l3-ifnlr_human-1,* and *l4-ifnlr_human-1.* Of these 19 genes, 4 are not present in the gene expression dataset. The venn diagram demonstrates overlap with Type I and Type II IFN gene sets used in the analyses shown in this study; of the 15 Type III genes present in the gene expression dataset, at least 3 genes are more classically associated with a Type II than a Type III IFN response (*IFNG, IFNGR1, IFNGR2*). **(B)** For each gene in the Type III IFN gene set, the percent of samples for which that gene had 0 detected counts was calculated, and shown for each anatomical site. Many samples demonstrated 0 counts for canonical Type III IFN genes (*IFNL1, IFNL2, IFNL3, IFNL4).* **(C)** The longitudinal expression of Type III IFN genes are shown for each anatomical site, with each line representing a participant. Lines colored black represent data from uninfected participants, and shown relative to enrollment in the study, while lines colored mauve represent data from participants with SARS-CoV-2 infection. **(D)** A module score was calculated using the Type III IFN gene set, and the distribution of module scores for samples from uninfected versus infected participants were compared. Point color represents SARS-CoV-2 viral load within that given sample. **(E)** Given the presents of canonically Type II IFN related genes in the Type III IFN gene set, Type II and Type III IFN module scores were compared for each anatomical site to assess for correlation.

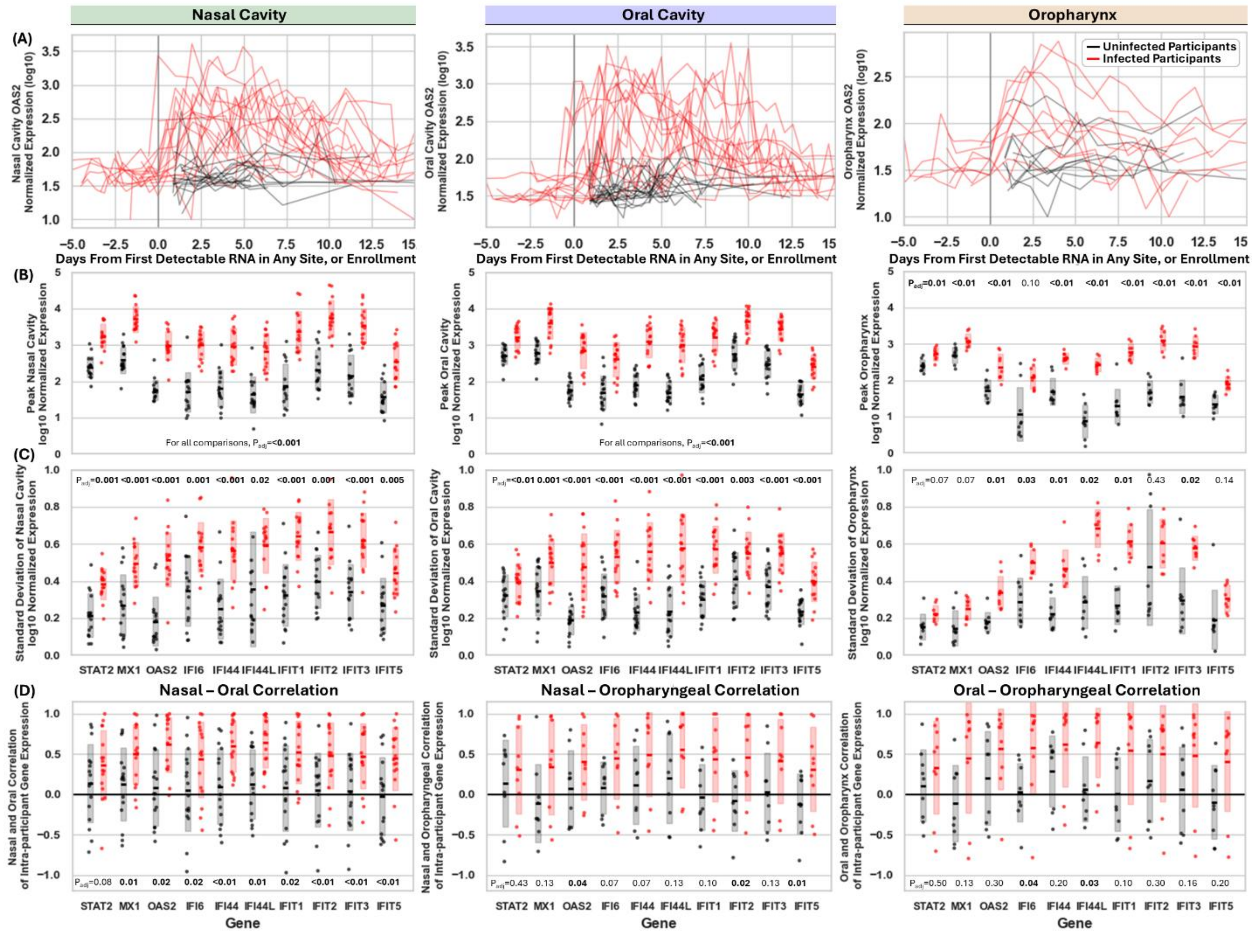


**Figure S4. Canonical Type I Interferon Stimulated Genes exhibits relatively low and stable expression within uninfected individuals, but high and dynamic expression in the nasal cavity, oral cavity, and oropharynx of infected individuals. (A)** Longitudinal normalized expression of OAS2 within uninfected participants (black lines) relative to time from enrollment, and within infected participants (red lines) relative to time from first detectable SARS-CoV-2 RNA in any anatomical site, for each anatomical site. A pseudocount of 10 was added to all datapoints, to enable log10 transformation including 2 of 483 nasal cavity datapoints and 3 of 233 oropharyngeal datapoints where OAS2 was not detected. **(B)** For a set of canonical Type I IFN Stimulated Genes, the peak normalized expression throughout the time course is shown for each uninfected participant (black points) and each infected participant (red points), for each anatomical site. Shaded region represents the standard deviation of log10 transformed values. **(C)** For a set of canonical Type I IFN Stimulated Genes, the standard deviation of log10-transformed normalized expression over time is shown for each uninfected participant (black points) and each infected participant (red points), for each anatomical site. Shaded region represents the participant-population standard deviation of individual-participant longitudinal standard deviation of expression. **(D)** For a set of canonical Type I IFN Stimulated Genes, pairwise correlation of normalized gene expression between anatomical sites within each uninfected participant (black points) and each infected participant (red points). Wilcoxon Rank Sum Test with Benjamini-Hochberg Correction was performed to test whether correlation coefficients were higher for infected participants than for uninfected participants; adjusted P values are shown above the corresponding gene on each panel.

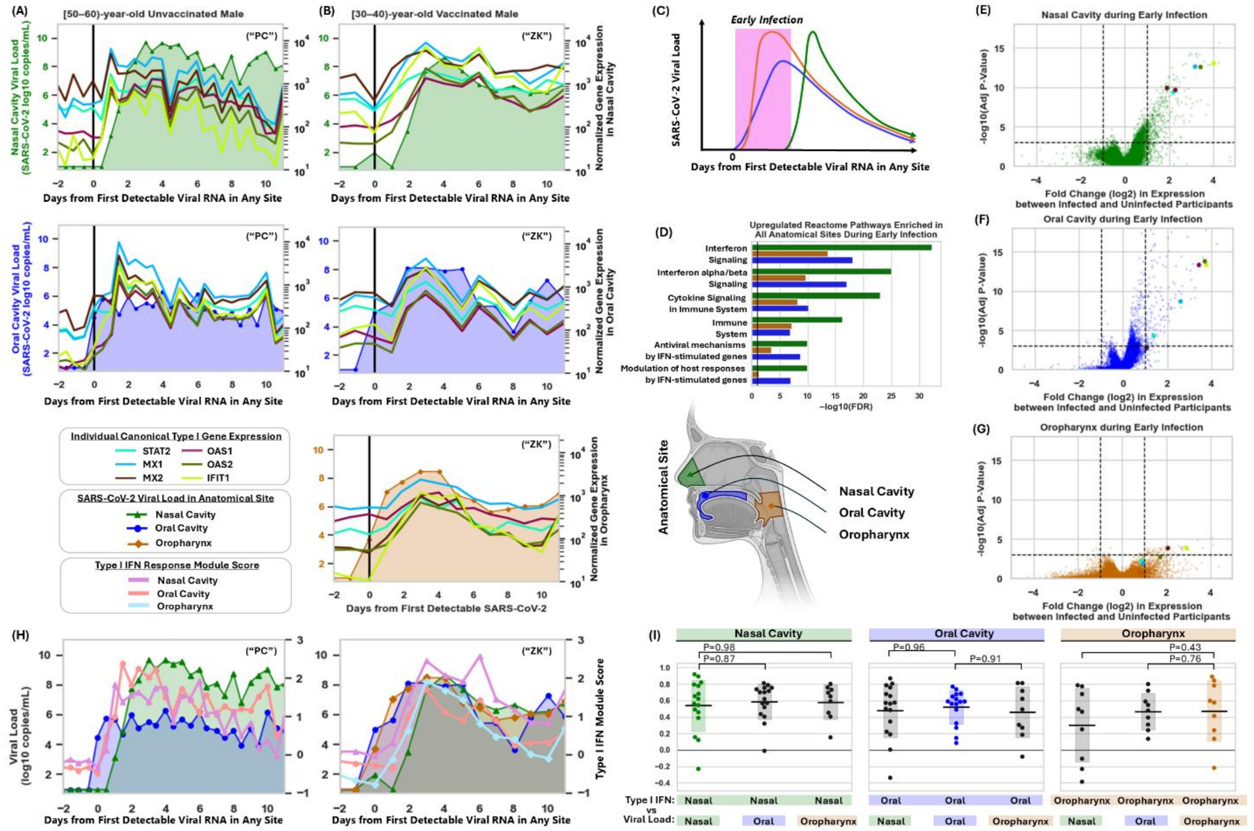


**Figure S5. Type I IFN-stimulated genes and a Type I IFN-predominant module increase concurrently across upper respiratory sites during early infection.** Canonical Type I IFN-stimulated gene expression is plotted with SARS-CoV-2 viral load over time at each anatomical site for a representative unvaccinated **(A)** and vaccinated **(B)** participant; "P" and "Z" identifiers refer to Participant ID as listed in **Table S1**. **(C)** Schematic of typical oral cavity, oropharyngeal, and nasal viral-load trajectories, to illustrate early infection (pink) as the interval from first detectable SARS-CoV-2 RNA at any site to first nasal detection, which was delayed in most infected participants. Differential expression between participants during early infection and uninfected participants by Mann–Whitney U test with Bonferroni correction. **(D)** The 100 most significantly upregulated genes per site were analyzed in DAVID and six ontology terms with lowest mean FDR are shown. **(E–G)** Volcano plots show differential expression results, with points indicating genes and selected Type I IFN-stimulated genes highlighted. Dashed lines indicate adjusted $P$ = 0.001 and fold change = ±2. **(H)** For the participants shown in **A** and **B**, expression of a previously described 25-gene Type I IFN-predominant program was summarized as a module score[5,11] and overlaid with local viral loads. Genes in this set include: *BST2, CMPK2, EIF2AK2, EPSTI1, HERC5, IFI35, IFI44L, IFI6, IFIT3, ISG15, LY6E, MX1, MX2, OAS1, OAS2, PARP9, PLSCR1, SAMD9, SAMD9L, SP110, STAT1, TRIM22, UBE2L6, XAF1,* and *IRF7*. **(I)** For each infected participant, longitudinal Type I IFN module scores were correlated with viral load either in the same site or other upper respiratory sites from that participant. Local versus cross-site correlation coefficients for each participant are compared pairwise by Mann–Whitney U tests without correction.

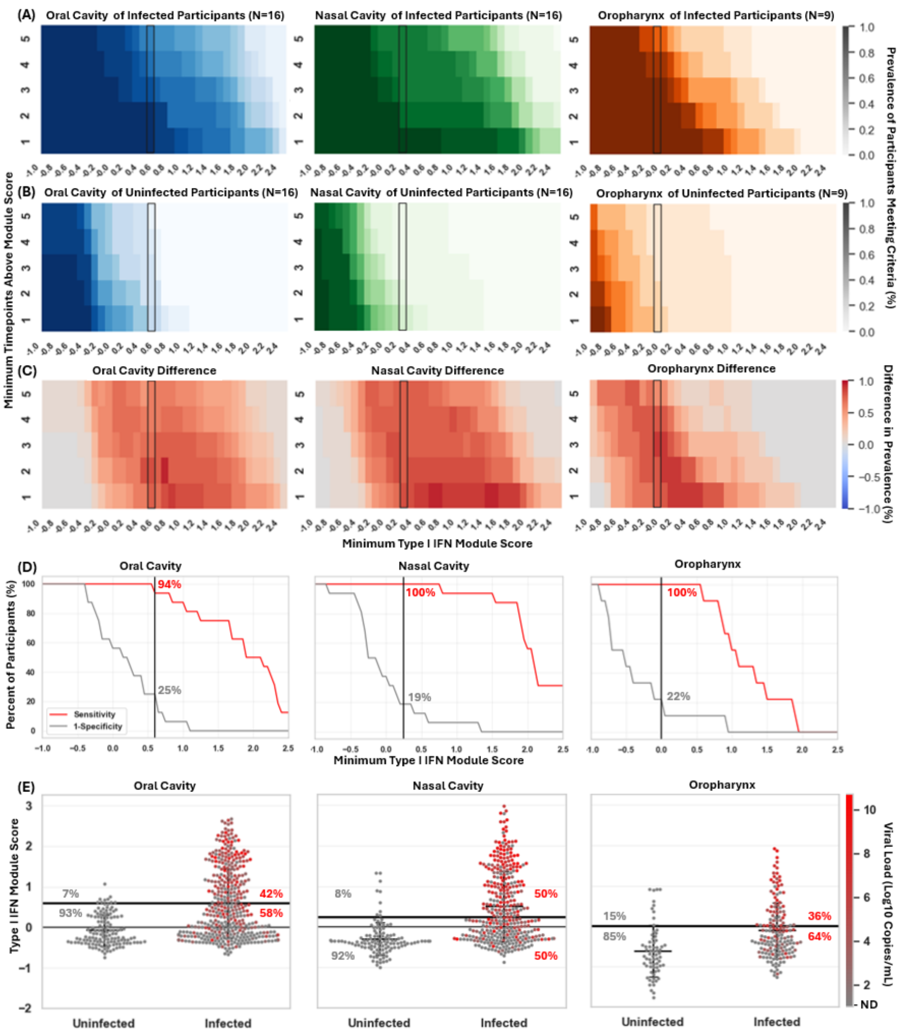


**Figure S6. Type I IFN-predominant response module scores in nasal cavity, oral cavity, and oropharyngeal samples differentiate uninfected from SARS-CoV-2 infected individuals.** For each sample, individual gene expression was used to calculate a module Score of expression for a set of 25 genes describing a predominantly Type I IFN response program, as previously described.[5,11] To assess the sensitivity and specificity of module scores for differentiating infection status in each anatomical site, the prevalence of participants - either infected **(A)** or uninfected **(B)** - with a minimum number of samples from that site (y-axis) above a minimum module score (x-axis) was calculated as the number of participants in group meeting criteria divided by all participants in the group. The difference between prevalence of infected participants meeting criteria minus the prevalence of uninfected participants meeting criteria **(C)** provides a visualization of how well a given module score threshold differentiates infected from uninfected participants. **(D)** For each anatomical site, a minimum threshold black was selected that achieved high (>90%) sensitivity and high (>75%) specificity for infection, based on at least one sample with Type I IFN Module Score greater than or equal to the threshold. Percentages indicate the proportion of infected (red) or uninfected (gray) participants meeting criteria, for this threshold. **(E)** In addition to differentiation by infection status, this threshold also differentiated samples by SARS-CoV-2 viral load (color). Percentages indicate the proportion of infected or uninfected samples above or below the threshold. Black box in **(A-C)**, black vertical line in **(D)**, and black horizontal line in **(E)** represent the selected threshold demarcating initiation of the Type I IFN response, in each anatomical site. Genes in this set include: *BST2, CMPK2, EIF2AK2, EPSTI1, HERC5, IFI35, IFI44L, IFI6, IFIT3, ISG15, LY6E, MX1, MX2, OAS1, OAS2, PARP9, PLSCR1, SAMD9, SAMD9L, SP110, STAT1, TRIM22, UBE2L6, XAF1,* and *IRF7*.

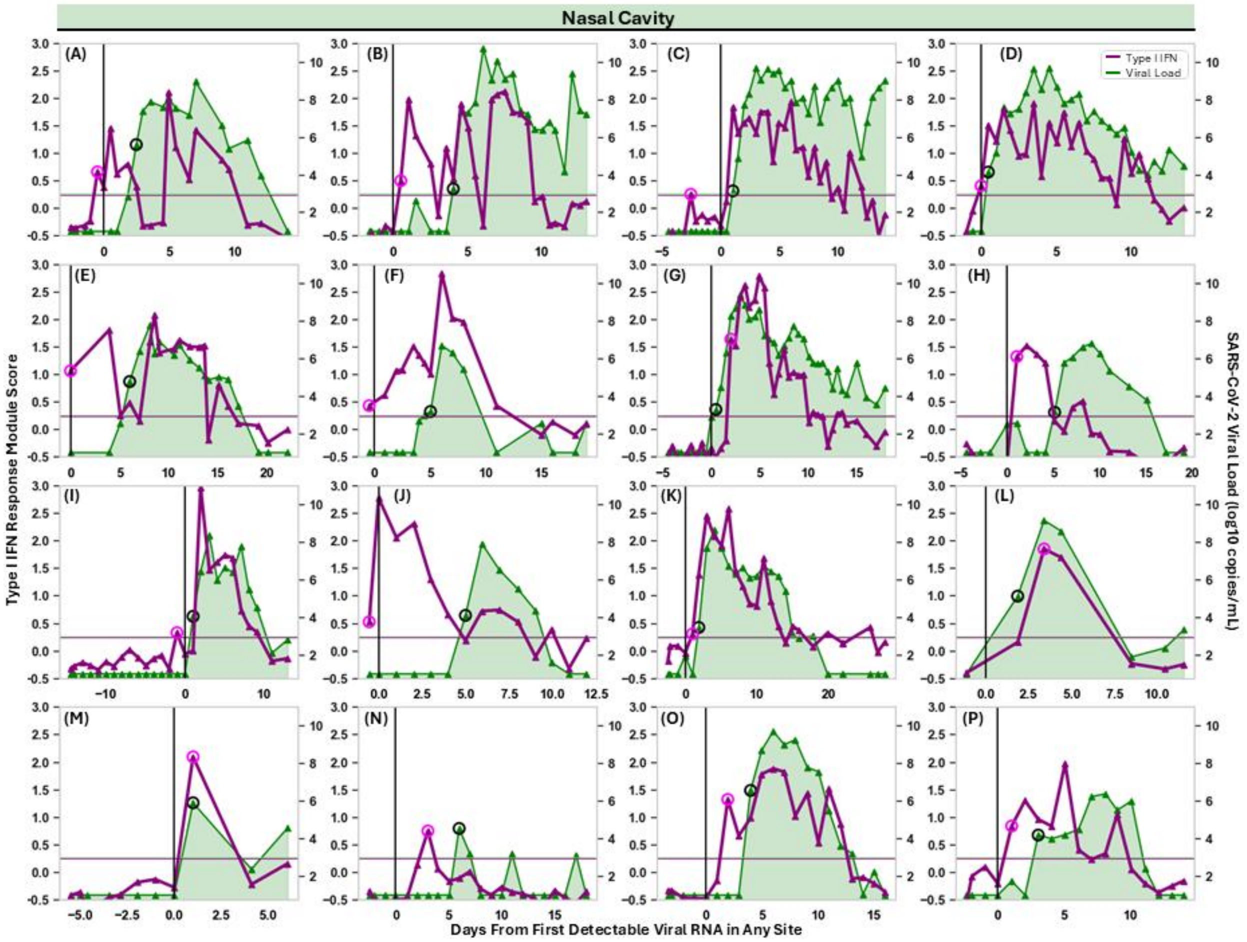


**Figure S7. Individual timeseries reveal a discernible interferon response preceding SARS-CoV-2 proliferation in the nasal cavity of infected individuals.** Type I IFN response module scores in the nasal cavity was plotted over time and overlayed with the local SARS-CoV-2 viral load from each infected participant **(A-P)**. Horizontal pink line indicates the threshold of module score which differentiates infected from uninfected participants (**Figure S3**), and horizontal gray line indicates a viral load of 1,000 copies/mL of specimen volume. The first timepoint to achieve a value above each of the threshold is indicated with a circle: a magenta circle represents the initiation of a discernible Type I interferon response in the nasal cavity, while a black circle represents the initiation of discernible viral proliferation in the nasal cavity. Panel letters correspond to "P" and "Z" identifiers as listed in **Table S1**.

***Alternate methods validate the synchronization of Type I IFN responses among upper respiratory mucosal sites during early SARS-CoV-2 infection***

Concurrent initiation of the Type I IFN response among upper respiratory anatomical sites within an individual was corroborated with two orthogonal analyses. Module scores based on an alternate Type I IFN gene set (MSigDB Hallmark IFNα Response, M5911),[12,13] were highly correlated with scores for the aforementioned Type I IFN gene set (**Figure S8**), and analyses using this alternate set yielded equivalent results.

We also performed single-sample gene set enrichment analysis (ssGSEA[4]) to score the magnitude of enrichment for KEGG Medicus pathways in each sample. Pathways associated with viral sensing and Type I IFN production (e.g. "MDA5 to IRF7/IRF3", "RIG-I to IRF7/IRF3", "TLR7/TLR9 to IRF7", "TRAF3-dependent IRF signaling") exhibited clear rises in all upper respiratory anatomical sites of infected participants during early infection, relative to low and stable expression among uninfected individuals (**Figure S9**). In contrast, the "TLR5 to NF-κB" signaling pathway, which was not expected to exhibit a strong response during early SARS-CoV-2 infection, did not appreciably rise among infected participants.

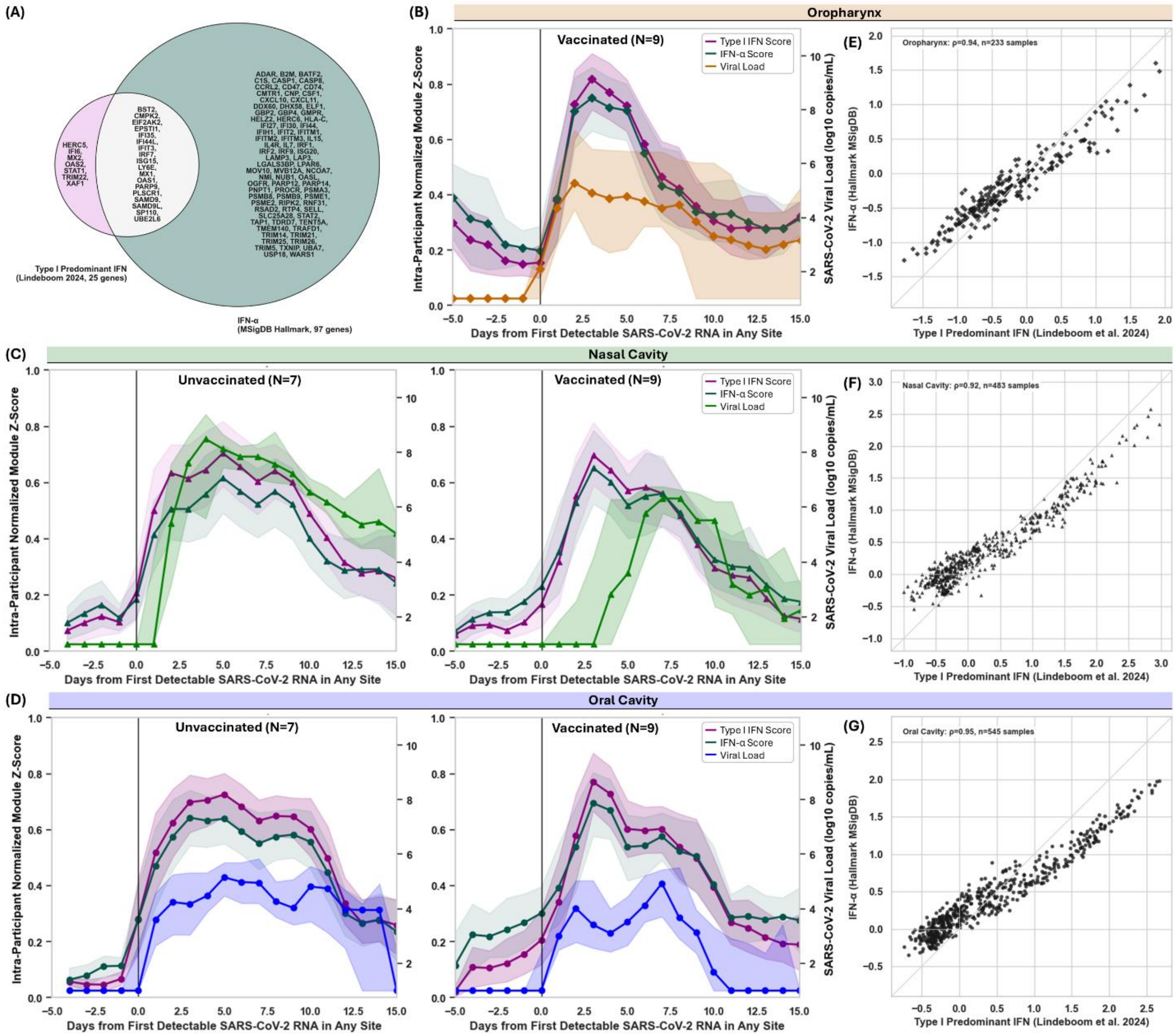


**Figure S8. Module scores for the Type I IFN predominant gene set match those resulting from a distinct IFN-α gene set from a separate source. (A)** Genes comprising two well-established, publicly available Type I IFN (IFNα/β) response gene sets, in pink[5,11] and the MSigDB IFNα Hallmark gene set (M5911, teal),[12,13] are compared in a Venn diagram. For each sample, module scores were computed using each gene set and normalized to scores (see Methods). Module scores were generated for each sample using these distinct gene sets. Module scores in samples from infected participants are overlaid against time from the beginning of infection in the **(B)** oropharynx, **(C)** nasal cavity, and **(D)** oral cavity, stratified by pre-infection COVID-19 vaccination status. Points indicate the mean module score across infected participants within 2-day time bins; shaded regions indicate 95% confidence intervals estimated by bootstrapping. **(E-G)** Module scores derived from the two gene sets were compared across all samples from both infected and uninfected participants in each anatomical site. Each point represents a single sample. Spearman correlation coefficients (ρ) are shown in each panel. Grey diagonal lines indicate the identity line.

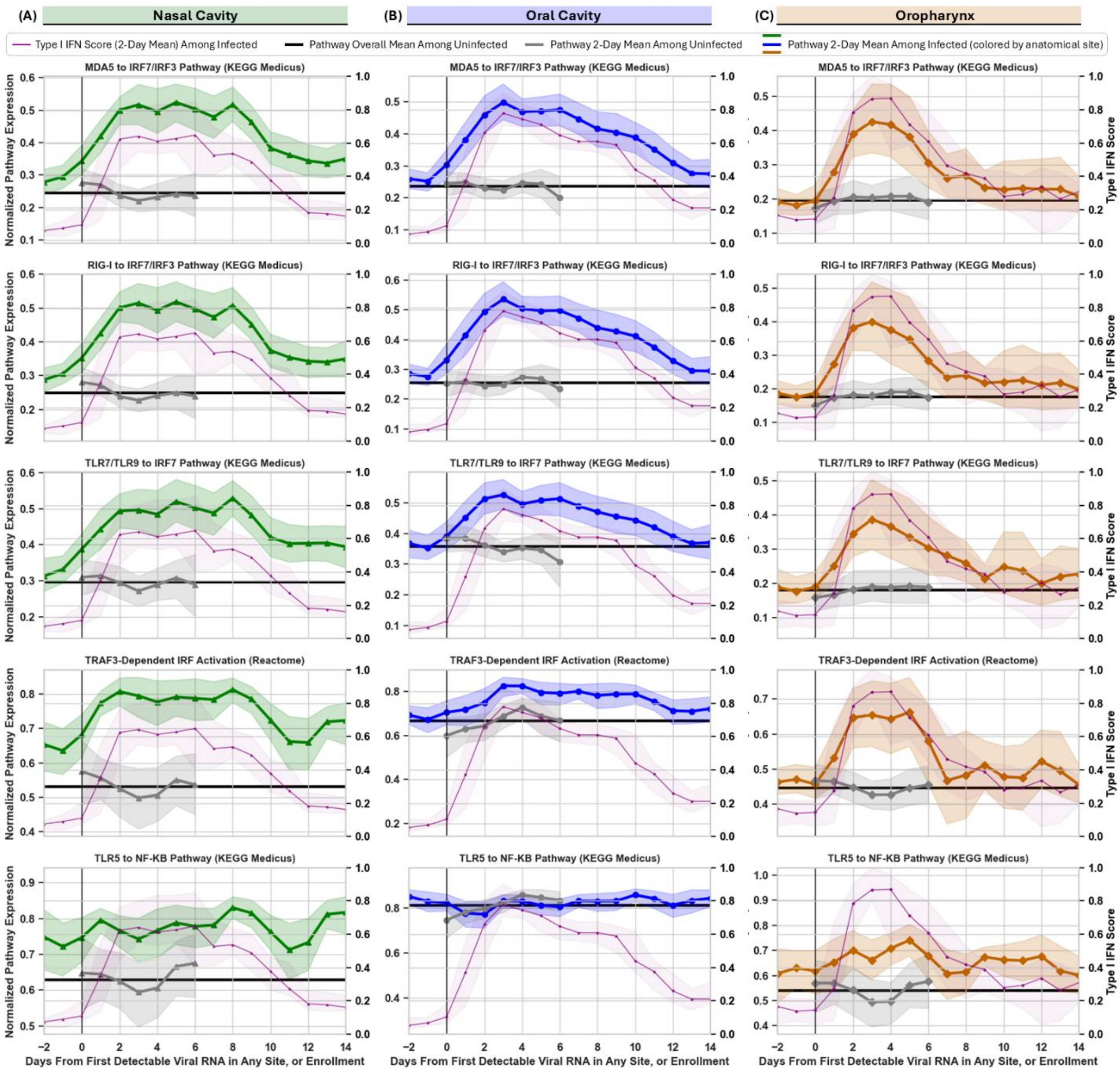


**Figure S9. Early activation of viral sensing pathways upstream of interferon production across upper respiratory anatomical sites during SARS-CoV-2 infection.** Single-sample Gene Set Enrichment Analysis (ssGSEA) was used to compute enrichment scores for curated KEGG Medicus pathways representing upstream viral sensing and signaling mechanisms, including viral sensing pathways “MDA5 to IRF7/IRF3”, “RIG-I to IRF7/IRF3”, “TLR7/9 to IRF7”, “TRAF3-dependent IRF activation”, and a bacterial flagellin sensing “TLR5 to NF-κB” signaling pathway. **(A-C)** For each anatomical site, within participant normalized scores were aggregated among participants by averaging rolling 2-day time bins relative to either the first timepoint with detectable SARS-CoV-2 RNA in any anatomical site for infected participants, or from enrollment for uninfected participants. Aggregate Type I IFN response module scores (reproduced from **Figure 1D**) for each anatomical site are overlayed. Shaded regions represent 95% confidence intervals estimated by bootstrapping. Vertical black lines indicate Day 0 for each group. Horizontal black lines indicate the mean pathway score across all samples from uninfected participants for that anatomical site.

***Activation of dendritic cells and macrophages likely mediate synchronized Type I IFN responses among upper respiratory mucosal anatomical sites of SARS-CoV-2 infected individuals***

To identify which cell types most likely contributed to the synchronous Type I IFN response in these upper respiratory mucosal sites, we performed cell type deconvolution on our gene expression data. Using a cell type signature matrix produced from scRNA-seq data generated from nasal cavity samples,[5] we found that the relative abundance of dendritic cells and macrophages was highly correlated with Type I IFN module scores in each site, for all three sites (**Figure S10A-C**). The abundance of these cell types increased during early infection, concomitant with the observed Type I IFN response (**Figure S10D-F**).

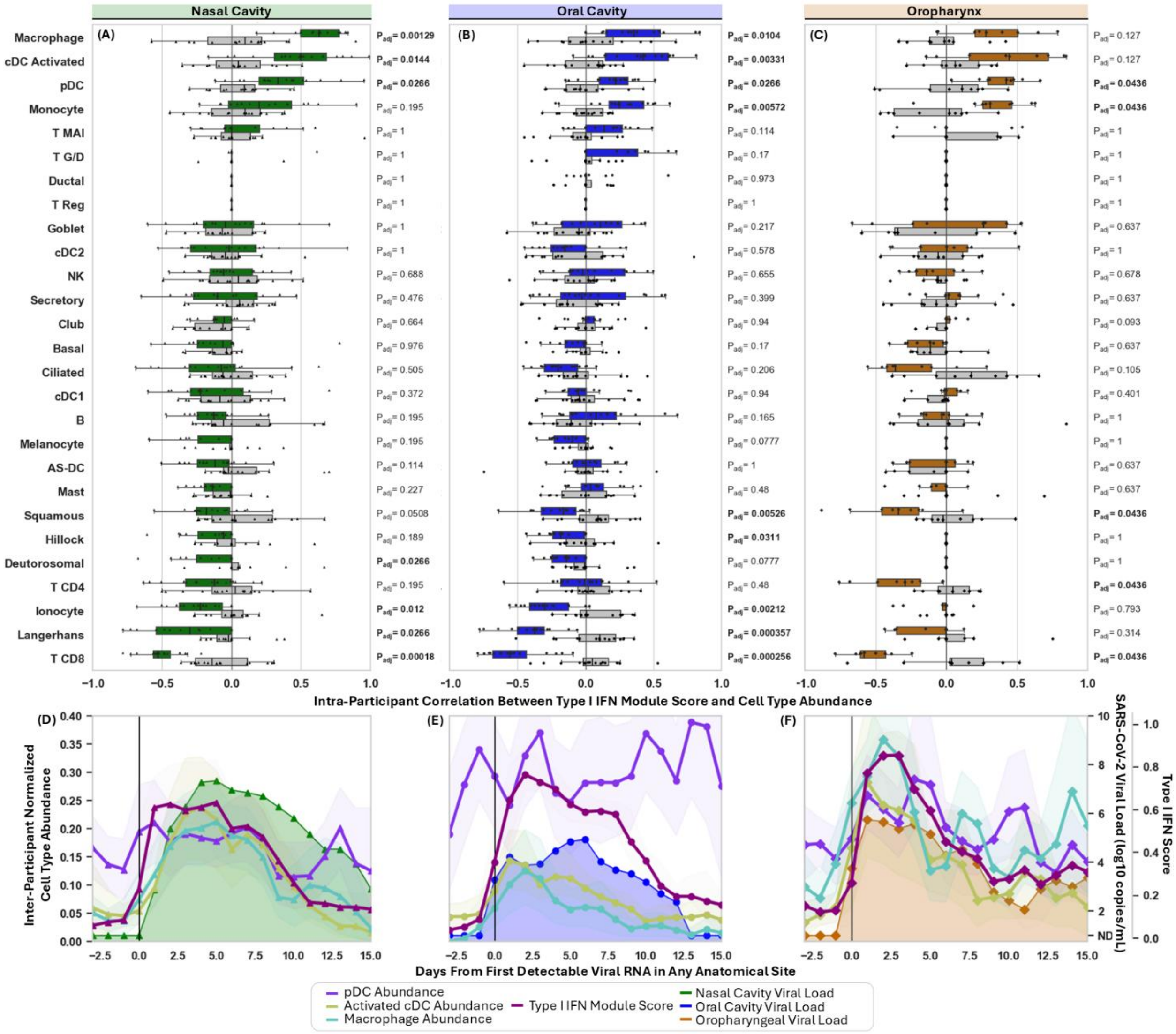


**Figure S10. Cell type abundance deconvolution suggests activated cDC and Macrophages promote the Type I IFN response in upper respiratory anatomical sites during acute SARS-CoV-2 infection.** For each anatomical sampling site **(A-C)**, the correlation between the Type I IFN Module Score and the relative abundance of each cell type over time within each individual with incident SARS-CoV-2 infection. Cell type abundances were calculated by deconvolution using CIBERSORTx and a Signature Matrix produced from scRNA-seq data reported in Lindeboom et al. 2024. Pearson correlation coefficient values for each participant were plotted and compared to a null model of coefficients resulting from time-shuffled samples for each participant using Wilcoxon Rank Sum Test with Benjamini Hochberg correction. **(D-F)** The longitudinal relative abundance of activated cDCs (gold) and macrophages (turqoise) were aggregated in 2-day time bins among participants (left y-axis), and plotted over time alongside SARS-CoV-2 viral load (internal right y-axis), and Type I IFN Module Score (external right y-axis).

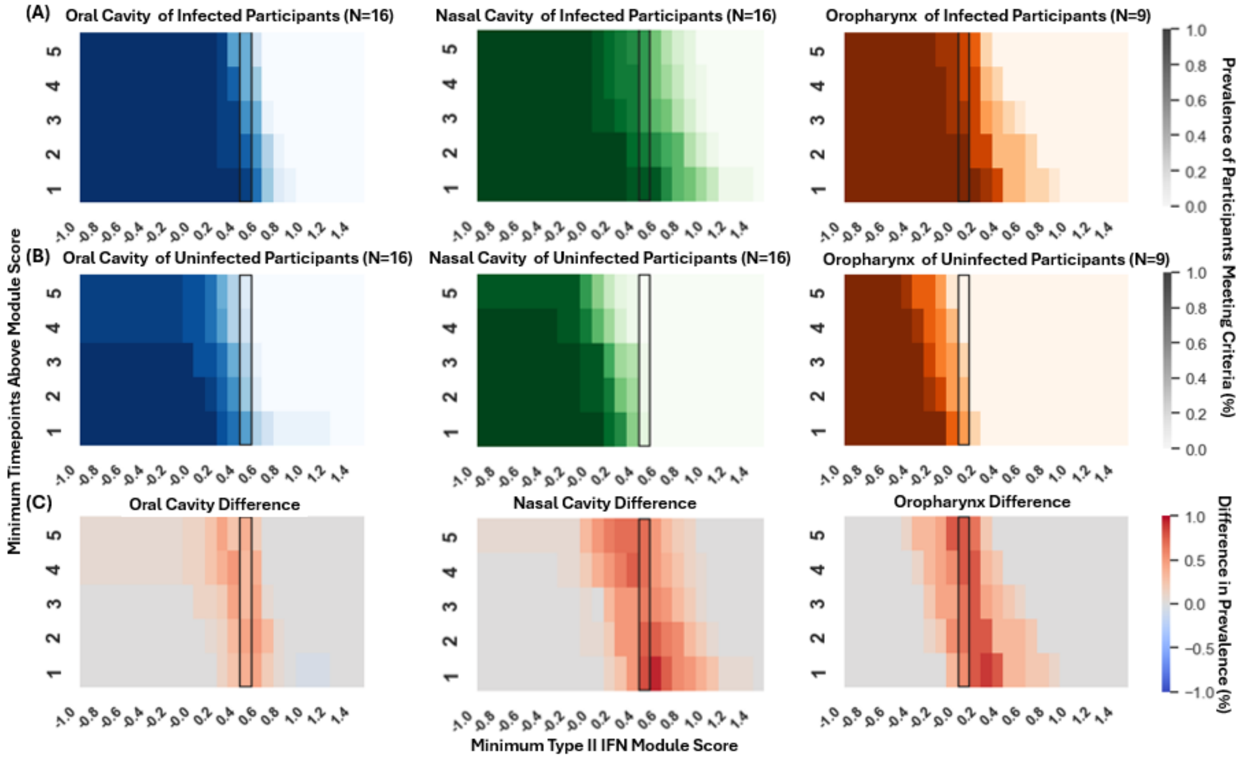


**Figure S11. Module Scores for a Type II Interferon Stimulated Gene Set differentiates uninfected from infected participants.** For each sample, individual gene expression was used to calculate a module Score of expression for a set of 64 genes describing a predominantly Type II IFN response program, listed as GO:0034341 (response to type II interferon, https://amigo.geneontology.org/amigo/term/GO:0034341). To assess the sensitivity and specificity of module scores for differentiating infection status in each anatomical site, the prevalence of participants – either infected **(A)** or uninfected **(B)** - with a minimum number of samples from that site (y-axis) above a minimum module score (x-axis) was calculated as the number of participants in group meeting criteria divided by all participants in the group. The difference between prevalence of infected participants meeting criteria minus the prevalence of uninfected participants meeting criteria **(C)** provides a visualization of how well a given module score threshold differentiates infected from uninfected participants. *ACOD1, ADAMTS13, AIF1, AQP4, ASS1, BST2, CALCOCO2, CALM1, CAMK2A, CASP1, CCL3, CD200, CD40, CDC42, CDC42EP4, CLDN1, CYP27B1, DAPK3, DNAJA3, EDN1, EPRS1, FLNB, GAPDH, GBP1, GBP2, GBP4, GBP5, GBP6, GBP7, GCH1, GPR146, GSN, HCK, HLA-DPA1, HPX, IFNG, IFNGR1, IFNGR2, IL12RB1, IL23R, KYNU, MEFV, NUB1, PDE12, PIM1, RAB43, RAB7B, RPL13A, SEC61A1, SHFL, SIRPA, SLC11A1, SLC22A5, SLC26A6, STAT1, TLR2, TLR3, TNF, TP53, UBD, VAMP4, VIM, WAS, WNT5A*

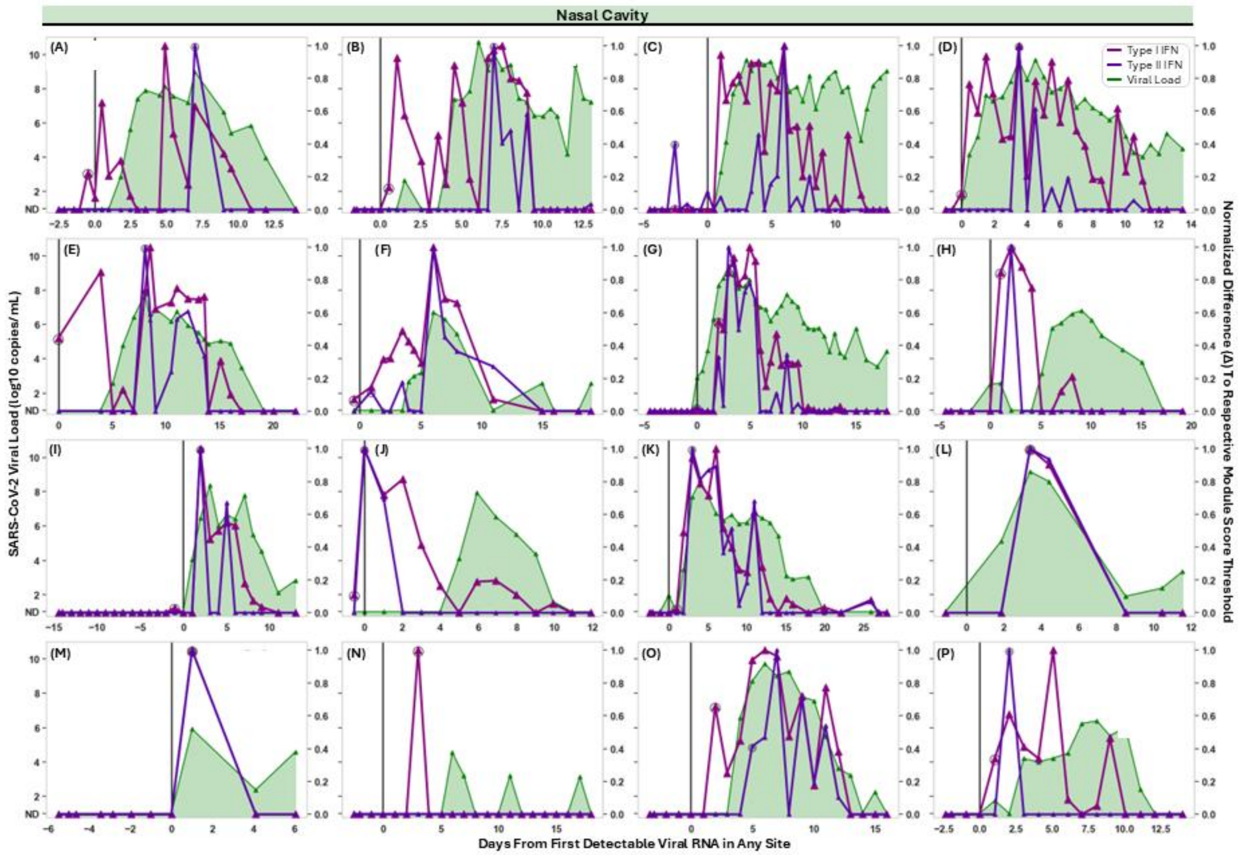


**Figure S12. Relative timing of Type I and Type II IFN Responses, and SARS-CoV-2 Viral Loads in the Nasal Cavities of Infected Individuals.** Type I IFN response module scores, previously described [https://doi.org/10.1038/s41586-024-07575-x, https://doi.org/10.1038/s41586-021-04345-x], and 64 genes describing a predominantly Type II IFN response program, listed as GO:0034341 (response to type II interferon)[10]. In the nasal cavity of each infected participant **(A-P)**, the intra-participant normalized Type I IFN module score (pink) and Type II IFN module score (purple) is plotted over time and overlayed with the SARS-CoV-2 viral load, with module scores clipped at the threshold differentiating infected from uninfected individuals (**Figure S6, Figure S11**). The first timepoint to achieve a value above each threshold is indicated with a circle. ND indicates viral RNA was not detected. Panel labels refer to "P" and "Z" participant identifiers listed in **Table S1**.

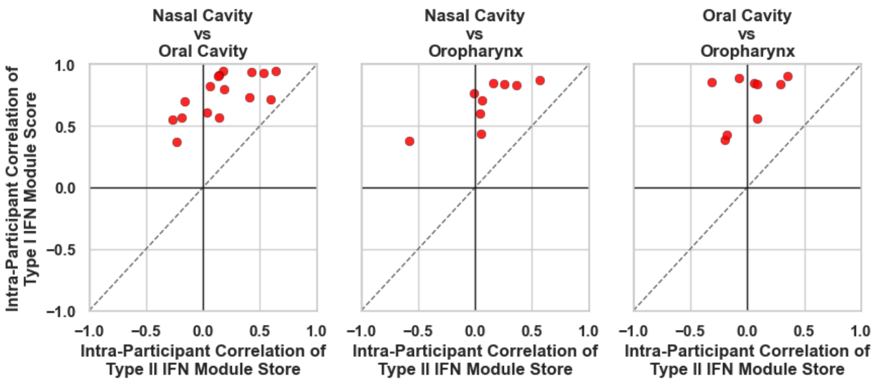


**Figure S13. The Type I IFN response correlates better among upper respiratory anatomical sites with an individual than the Type II IFN response.** Apart from synchronization in the initiation of Type I or Type II response timing, the correlation of the response among upper respiratory sites was analyzed. For each participant (datapoint), the correlation of Type I IFN module score between pairwise upper respiratory sites is plotted (y-axis) against the correlation of Type II IFN module scores (x-axis) between these sites. Datapoints above the identity line indicate that Type I IFN correlates better among upper respiratory sites than Type II IFN.

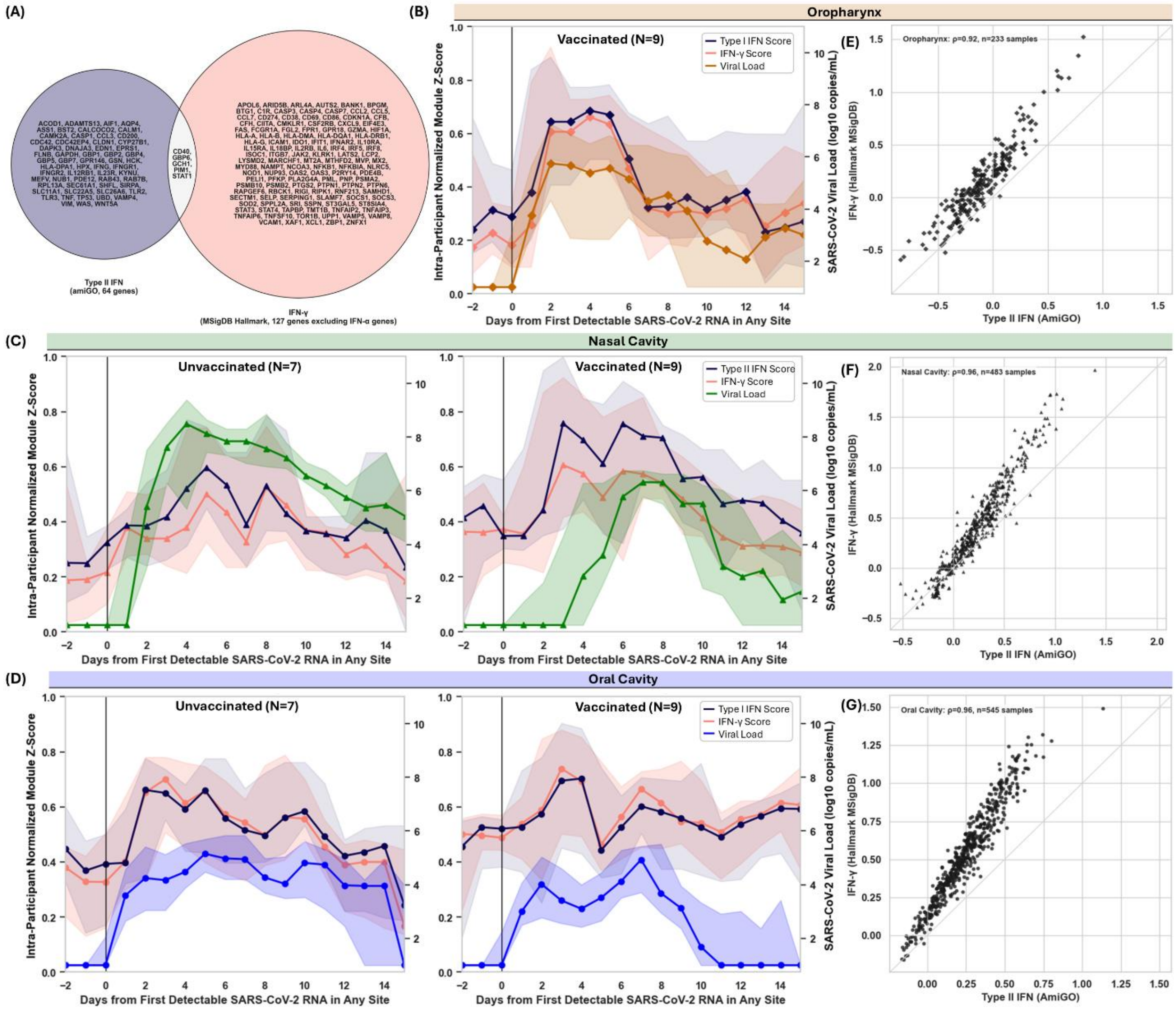


**Figure S14. Module scores for the Type II IFN gene set match those resulting from a distinct IFNγ gene set from a separate source.** Genes comprising two well-established, publicly available Type II IFN (IFNγ) response gene sets - AmiGO Type II IFN Response (GO:0034341, orchid) the MSigDB IFNγ Hallmark gene set (M5913,[12,14] salmon) - are compared in **(A)** a Venn diagram. For each sample, module scores were computed using each gene set and normalized to scores (see **Methods**). Module scores were generated for each sample using these distinct gene sets. Module scores in samples from infected participants are overlaid against time from the beginning of infection in each anatomical site: oropharynx **(B,C)**, nasal cavity **(E,F)**, and oral cavity **(H,I)**, and stratified by participant vaccination status. Points indicate the mean module score across infected participants within 2-day time bins; shaded regions indicate 95% confidence intervals estimated by bootstrapping. Module scores derived from the two gene sets were compared directly across all samples from both infected and uninfected participants in the oropharynx **(D)**, nasal cavity **(G)**, and oral cavity **(J)**. Each point represents a single sample. Spearman correlation coefficients (ρ) are shown in each panel. Grey diagonal lines indicate the identity line.

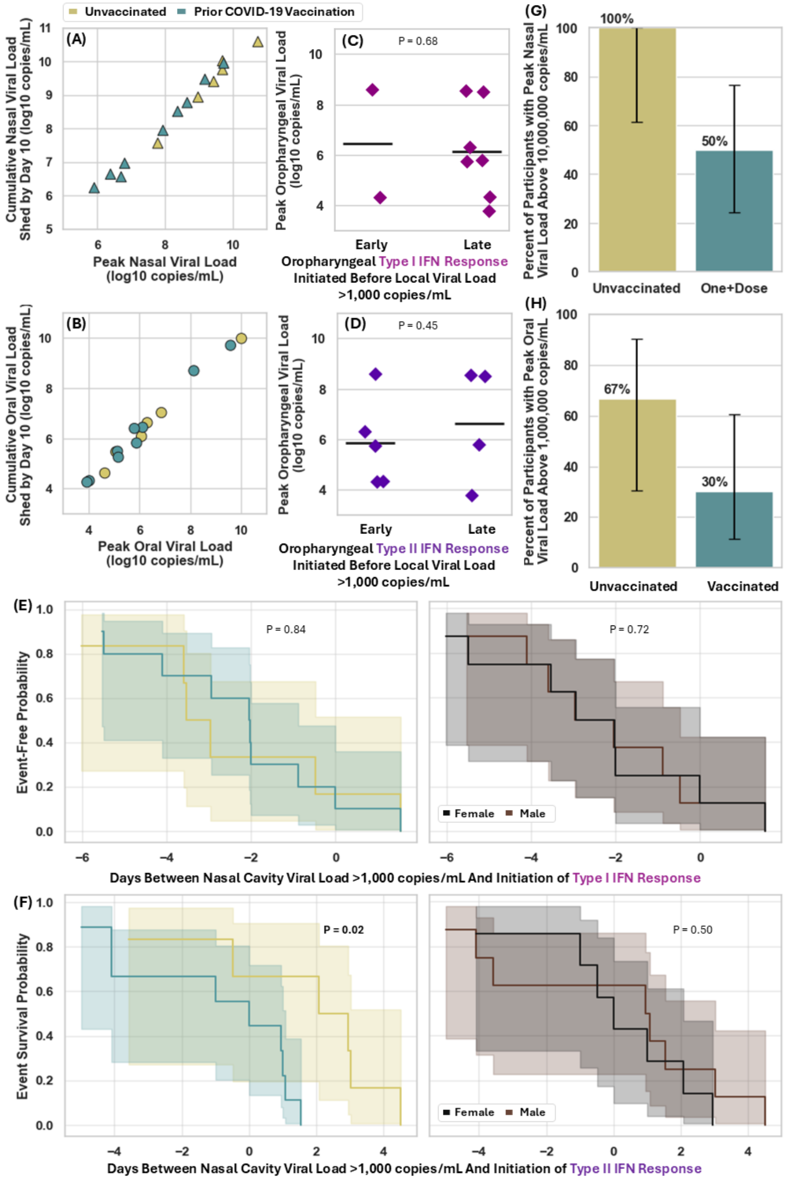


**Figure S15. Additional analyses of the interplay between vaccination, Type I IFN, Type II IFN and viral load within upper respiratory anatomical sites.** In the nasal cavity **(A)** and the oral cavity **(B)**, peak viral load for each participant was plotted against cumulative virus shed by day 10 of infection for that participant, calculated as the area under curve of daily viral load measurements, colored by vaccination status. **(C-D)** Participants were classified as exhibiting early versus late initiation of Type I **(C)** or Type II **(D)** IFN responses in the oropharynx based on whether a module score differentiable from uninfected participants occurred prior to viral load of $10^3$ copies/mL in the oropharynx. Distributions of peak oropharyngeal viral load were compared between participants with early versus late IFN response initiation of by Wilcoxon Rank Sum Test. In the nasal cavity, time-to-event analyses were performed using viral load above 1,000 copies/mL and initiation of the Type I IFN response **(E)** or Type II IFN response **(F)**, stratified by either prior COVID-19 vaccination (left) or participant sex (right). P-value calculated by Log-rank test. In the nasal cavity **(G)** and oral cavity **(H)** the percent of infected participants with presumably infectious peak viral loads (above 10,000,000 or 1,000,000 copies/mL respectively) are shown, stratified by prior COVID-19 vaccination. Error bars represent binomial error.

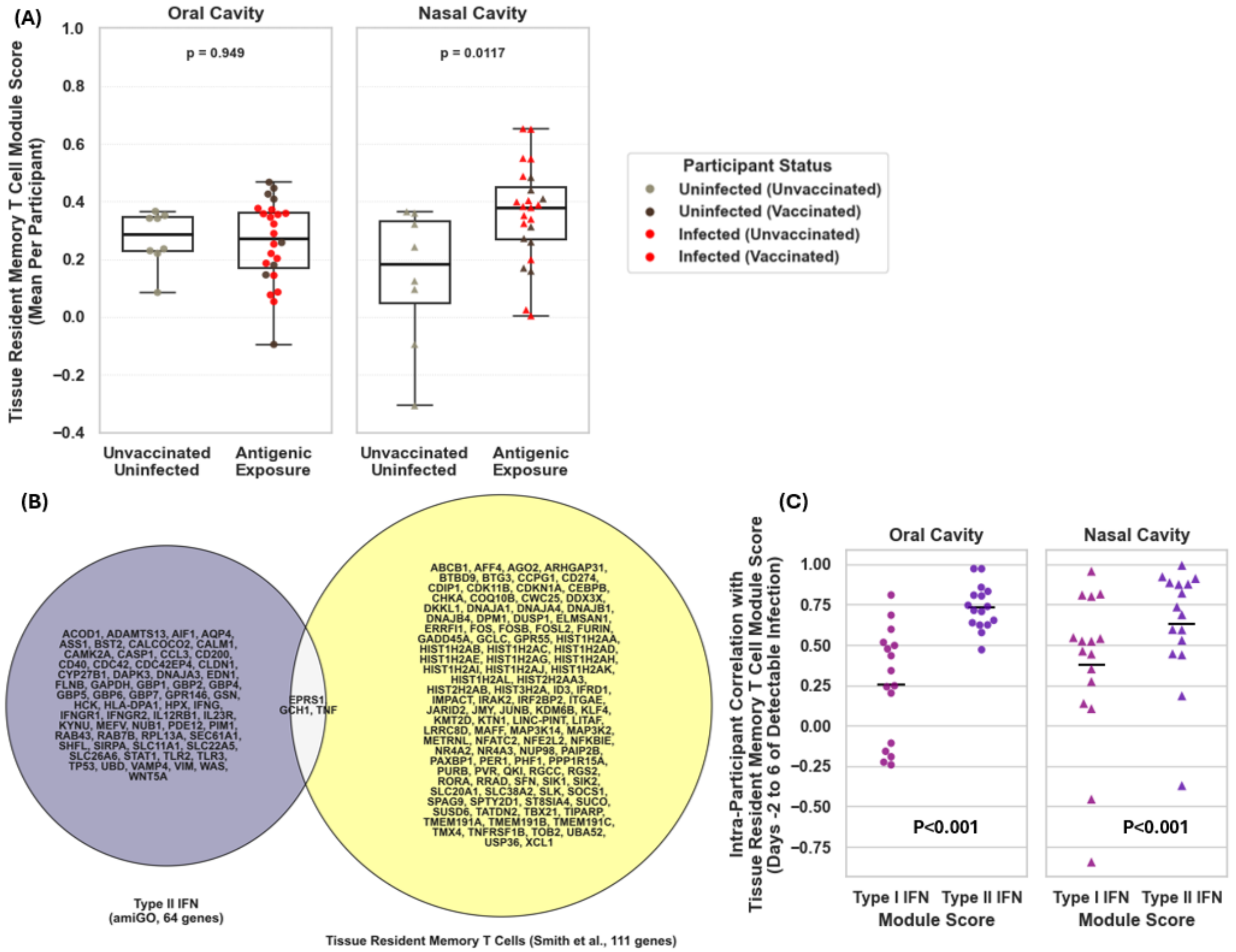


**Figure S16. Additional analyses of SARS-CoV-2 antigenic exposure, tissue resident memory T cell scores, and Type I and Type II IFN responses.** A module Score for tissue resident memory T cells in each sample were calculated using a previously defined set of 112 genes.[15] **(A)** Participants with SARS-CoV-2 antigenic exposure due to sustained incident infection (regardless of vaccination status) or due to parenteral vaccination (regardless of infection status) were grouped, and compared to participants without antigenic exposure (unvaccinated, uninfected individuals). The mean $T_{RM}$ module score was calculated among all samples from a given anatomical sampling site from each participant, and compared by Mann Whitney U Test. **(B)** The gene set comprising the tissue resident memory T cell ($T_{RM}$) module score was compared to the gene set comprising the Type II IFN response module score. **(C)** For each infected participant, the correlation between $T_{RM}$ module score and either Type I or Type II IFN response module score was calculated. The distribution of these intra-participant correlation coefficients were plotted. The distribution of correlation coefficients for Type I IFN response module scores were compared to the correlation coefficients for Type II IFN response module scores, for each anatomical site, using Wilcoxon Signed Rank Test.

**Table S2. Summary of published studies describing human mucosal responses during SARS-CoV-2 infection.** Relevant literature describing human mucosal immune responses during acute SARS-CoV-2 infection, in either single or multiple anatomical sites, in a cross-sectional or longitudinal manner, and during incident infection or after symptom onset.

| Citation | Infection Stage | Mucosal Immune Site(s) | Temporal Kinetics | Viral Variant |
|---|---|---|---|---|
| Lindeboom, R.G.H., Worlock, K.B., Dratva, L.M. et al. Human SARS-CoV-2 challenge uncovers local and systemic response dynamics. Nature **631**, 189–198 (2024). | Incident | Nasopharynx | Longitudinal | Wild-type Pre-Alpha |
| Rosenheim, J., Gupta, R.K., Thakker, C. et al. SARS-CoV-2 human challenge reveals biomarkers that discriminate early and late phases of respiratory viral infections. Nat Commun **15**, 10434 (2024). | Incident | Nose | Longitudinal | Wild-Type Pre-Alpha |
| Ramirez, S.I., Faraji, F., Hills, L.B. et al. Immunological memory diversity in the human upper airway. Nature **632**, 630–636 (2024). | Post-infection | Nasopharynx, Nasal turbinate, Adenoid | Longitudinal | Not reported |
| Yoshida, M., Worlock, K.B., Huang, N. et al. Local and systemic responses to SARS-CoV-2 infection in children and adults. Nature **602**, 321–327 (2022). | Following symptom onset, convalescence | Nose, Trachea, Bronchi | Cross-sectional during infection, additional convalescent timepoint | Not reported, before March 2021 |
| Mick, E., Kamm, J., Pisco, A.O. et al. Upper airway gene expression reveals suppressed immune responses to SARS-CoV-2 compared with other respiratory viruses. Nat Commun **11**, 5854 (2020). | Following symptom onset | Nasopharynx, Oropharynx | Cross-sectional | Ancestral, March and April 2020 |
| Dianna L. Ng et al., A diagnostic host response biosignature for COVID-19 from RNA profiling of nasal swabs and blood. Sci. Adv. **7**, eabe5984 (2021) | Following symptom onset | Nasopharynx | Cross-sectional | Ancestral, before May 2020 |
| Gao, Kevin M et al. "Human nasal wash RNA-Seq reveals distinct cell-specific innate immune responses in influenza versus SARS-CoV-2." JCI insight vol. 6,22 e152288. 22 Nov. 2021 | Following symptom onset | Nasal Wash | Cross-sectional | Ancestral, before July 2020 |
| Islam, A.B.M.M.K., Khan, M.AAK., Ahmed, R. et al. Transcriptome of nasopharyngeal samples from COVID-19 patients and a comparative analysis with other SARS-CoV-2 infection models reveal disparate host responses against SARS-CoV-2. J Transl Med **19**, 32 (2021). | Following symptom onset | Nasopharynx | Cross-sectional | Not reported |
| Ziegler, Carly G K et al. "Impaired local intrinsic immunity to SARS-CoV-2 infection in severe COVID-19." Cell vol. 184,18 (2021) | Following symptom onset | Nasopharynx | Cross-sectional | Not reported, before Oct 2020 |
| Loske, J., Röhmel, J., Lukassen, S. et al. Pre-activated antiviral innate immunity in the upper airways controls early SARS-CoV-2 infection in children. Nat Biotechnol **40**, 319–324 (2022) | Following symptom onset | Nasopharynx | Cross-sectional | Not reported, before July 2021 |
| Mick, E., Tsitsiklis, A., Spottiswoode, N. et al. Upper airway gene expression shows a more robust adaptive immune response to SARS-CoV-2 in children. Nat Commun **13**, 3937 (2022). | Following symptom onset | Nasopharynx | Cross-sectional | Ancestral, pre Oct. 2020 |
| Rajagopala SV, Strickland BA,Pakala SB, et al. 2023. Mucosal Gene Expression in Response to SARS-CoV-2 Is Associated with Viral Load. J Virol 97:e01478-22. | Following diagnosis | Nasopharynx | Cross-sectional | Ancestral, Spring 2020 |
| Moradi Marjaneh, Mahdi et al. "Analysis of blood and nasal epithelial transcriptomes to identify mechanisms associated with control of SARS-CoV-2 viral load in the upper respiratory tract." The Journal of infection vol. 87,6 (2023): 538-550. | Following symptom onset | Nasopharynx | Cross-sectional | Ancestral, before June 2020 |
| https://journals.plos.org/plosone/article?id=10.1371/journal.pone.0317033 | Following symptom onset | Nasal mid-turbinate | Cross-sectional | Not reported, Mar 2020- Jan 2021 |
| Hurst, J.H., Mohan, A.A., Dalapati, T. et al. Age-associated differences in mucosal and systemic host responses to SARS-CoV-2 infection. Nat Commun **16**, 2383 (2025). | Following symptom onset | Nasopharynx | Cross-sectional | Ancestral, pre Jan. 2021 |
| Silva, Julio et al. "Saliva viral load is a dynamic unifying correlate of COVID-19 severity and mortality." medRxiv 2021 | Following symptom onset | Saliva, Nasopharynx | Cross-sectional | Ancestral, before July 2020 |
| Gómez-Carballa A, Rivero-Calle I, Pardo-Seco J, et al. A multi-tissue study of immune gene expression profiling highlights the key role of the nasal epithelium in COVID-19 severity. Environ Res. 2022 | Following symptom onset | Saliva, Nasal epithelium | Cross-sectional | Ancestral, before July 2020 |

### CRediT AUTHOR CONTRIBUTIONS

**Alexander Viloria Winnett:** Conceptualization, Methodology, Formal analysis, Investigation, Software, Visualization, Validation, Data curation, Project administration, Funding acquisition, Writing - Original Draft, Writing - Review & Editing. **Alexandra Tabachnikova:** Formal analysis, Validation, Data curation, Visualization, Writing - Original Draft, Writing - Review & Editing. **Jonathan Chen:** Investigation, Formal analysis, Validation, Data curation, Methodology, Writing - Review & Editing. **Kerrie Greene:** Formal analysis, Methodology, Writing - Review & Editing. **Anna E Romano:** Methodology, Investigation, Validation, Data curation, Writing - Review & Editing. **Xinyue Penny Pei:** Investigation, Validation, Data curation, Writing - Review & Editing. **Matthew M Cooper:** Methodology, Formal analysis, Visualization, Writing - Review & Editing. **Julio Silva:** Methodology, Formal analysis, Writing - Review & Editing. **Alyssa M Carter:** Investigation, Writing - Review & Editing. **Jialong Jiang:** Methodology, Software, Writing - Review & Editing. **Yong Kong:** Methodology, Software, Writing - Review & Editing. **Morgan Roos:** Methodology, Investigation, Writing - Review & Editing. **Christina Middle:** Methodology, Investigation, Writing - Review & Editing. **Hanqiao Zhang:** Formal analysis, Writing - Review & Editing. **Matt Thomson:** Methodology, Writing - Review & Editing. **Keith Booher:** Supervision, Resources, Writing - Review & Editing. **Scott Kuersten:** Methodology, Supervision, Resources, Writing - Review & Editing. **Akiko Iwasaki:** Conceptualization, Resources, Funding acquisition, Supervision, Writing - Original Draft, Writing - Review & Editing. **Rustem F Ismagilov:** Conceptualization, Resources, Project administration, Supervision, Funding acquisition, Writing – Original Draft, Writing - Review & Editing.

### DETAILED AUTHOR CONTRIBUTIONS

AVW - Conceptualized and designed study. Designed study budget with AER, and funding acquisition with RFI and AI. Established and maintained collaborations. Developed and validated a robust sample processing pipeline to generate high quality RNA sequencing data from complex upper respiratory mucosal clinical samples, with support from AER, SK, MR, CM, XPP and RFI. Assisted JC and KB in the conversion of this workflow to high-throughput, automated format, with metrics to monitor and algorithms to ensure RNA sequencing data quality control during generation. Developed the computational pipeline to process raw RNA sequencing data and generate gene expression data with support from HZ. Evaluated gene expression data quality and performed quality control analyses in Figure S1. Performed underlying data validation with XPP. Performed analyses of gene expression data, including development and validation of analytical methods with major support in development, validation and interpretation of results from AT, RFI and AI, as well as support from KG, JS, YK, JJ, MT, MMC, and AER. Performed analyses shown in Figure 1, Figure 2, Figure 3, Figure 4, Figure S1, Figure S2, Figure S3, Figure S4, Figure S5, Figure S6, Figure S7, Figure S8, Figure S9, Figure S10, Figure S11, Figure S12, Figure S13, Figure S14, Figure S15, Figure S16. Created Graphical Abstract with AT. Prepared Table S1. Outlined and drafted manuscript. Revised manuscript with AT, AI, and RFI, and co-authors. Coordinated and incorporated co-author feedback.

AT - Refined study design with AVW, RFI, AI, KG, and JS. Provided feedback on gene expression data normalization methods, specimen selection, case-control matching, cell type abundance estimation, and identification/validation of IFN and $T_{RM}$ gene sets. Major contributor to development of analyses resulting in Figure 1, Figure 2, Figure 3, Figure 4, Figure 5. Compiled Table S2. Created Graphical Abstract with AVW. Revised outline and drafted manuscript with AVW. Contributed to the writing and editing the manuscript.

JC - Major contributor to data generation. Developed and validated the high-throughput workflow for generating RNA sequencing data. Validated the automated RNA purification using Quick-DNA/RNA HT on KingFisher Flex. Based on foundational work from SK, validated the high throughput library preparation and enrichment procedure using the Illumina RNA Prep and Enrichment protocol. Developed quality control methods and metrics for each checkpoint in the workflow with approval from AVW and AER. Discussed and implemented workflow optimizations to generate data for a subset of difficult (low human biomass, high microbial background) samples with SK, AVW and KB. Oversaw and coordinated the sample processing and data generation from RNA extraction to sequencing. Provided significant feedback on manuscript draft.

KG - Refined study design with AVW, RFI, AI, AT, and JS. Provided feedback to AVW on computational RNA sequencing data processing pipeline, on gene expression data normalization methods, on specimen selection and on case-control matching. Provided significant feedback on manuscript draft.

AER - Designed study budget with AVW. Assisted in the development of robust sample processing pipeline to generate high quality RNA sequencing data from complex upper respiratory mucosal clinical samples. Evaluated and defined metrics to monitor and ensure RNA sequencing data quality control with AVW. Provided significant feedback on manuscript draft.

XPP - Developed a high-throughput DNA Qubit assay to assist AVW in the evaluation of candidate sample preparation methods for RNA sequencing, developed and implemented three-pass check system with AVW to ensure specimen aliquoting accuracy, and assisted AVW with aliquoting volume from >1200 BSL2 primary specimens. Major contributor to metadata validation (including compilation of several hundred viral load measurements). Provided feedback on manuscript draft.

MMC - Analyzed gene expression data to optimize methods for data normalization (specifically, handling of low abundance genes) and downstream analyses. Performed differential expression analysis and principal component analysis shown in Figure S3. Provided feedback on manuscript draft.

JS - Refined study design with AVW, RFI, AI, AT, and KG. Assisted AVW with interpretation of cell type abundance analyses. Provided feedback to optimize data normalization (specifically, TMM normalization), as well as identification and validation of IFN gene sets with AVW and AT. Provided feedback on manuscript draft.

AMC - Assisted AVW in early evaluation and optimization of sample preparation, as well as preparing samples for shipment to collaborators for validation experiments. Provided feedback on manuscript draft.

JJ - Provided feedback to AVW and HW to optimize methods for gene expression data normalization and downstream analyses (specifically, mean centering and variance stabilization). Provided feedback on manuscript draft.

YK - Assisted AVW in validation of RNA sequencing data quality. Assisted AVW in the development of RNA sequencing data processing methods, to generate gene expression data. Generated signature matrices using scRNA-seq data from which AVW performed cell type deconvolution and downstream analysis. Provided feedback on manuscript draft.

MR - Assisted with the development and validation of the technical performance of RNA sequencing data generation pipeline (specifically, optimized method for RNA sequencing library preparation). Provided feedback on manuscript draft.

CM - Assisted with the development and validation of the technical performance of RNA sequencing data generation pipeline (specifically, method for RNA sequencing library preparation). Provided feedback on manuscript draft.

HZ - assisted in the quality control of raw RNA sequencing data and development of RNA sequencing data processing pipeline to generate gene expression matrix with AVW. Provided feedback on manuscript draft.

MT - helped define sequencing technical specifications, sequencing data quality control, gene expression data normalization, feedback on downstream analyses. Provided feedback on manuscript draft.

KB - Oversaw conversion of sample preparation methods to high-throughput, automated format, and subsequent implementation of methods for high quality RNA sequencing data generation. Provided feedback on manuscript draft.

SK - Major contributor to the development and validation of library preparation, human exome enrichment, and RNA sequencing methods, including for difficult specimens (i.e. low human biomass, high microbial background). Provided feedback to JC and KB to convert methods to high-throughput automated format. Provided feedback on manuscript draft.

AI - Refined study design with AVW, RFI, AT, KG, and JS. Supported funding acquisition. Contributed to the writing, editing and refining the key message of the manuscript.

RFI - Conceptualized study with AVW. Funding acquisition. Study administration. Advised AVW on evaluation and optimization of sample processing methods for high quality, RNA sequencing data generation. Contributed to the writing and editing of the manuscript.

### AUTHOR CONTACT INFORMATION

| Name | Email | Phone (+1) | ORCiD |
|---|---|---|---|
| Alexander V Winnett | | 781-985-1502 | 0000-0002-7338-5605 |
| Alexandra Tabachnikova | | 203-785-7662 | 0000-0003-4695-2480 |
| Jonathan Chen | | 888-882-9682 | |
| Kerrie Greene | | 203-785-7662 | 0000-0002-2988-2436 |
| Anna E Romano | | 626-395-3464 | 0000-0002-7148-0668 |
| Xinyue Penny Pei | | 626-395-3464 | 0009-0003-9840-6243 |
| Matthew M Cooper | | 626-395-3464 | 0000-0002-5868-5159 |
| Julio Silva | | 203-785-7662 | 0000-0001-8212-7440 |
| Alyssa M. Carter | | 625-722-7526 | 0000-0002-2776-9421 |
| Jialong Jiang | | 626-395-8782 | 0000-0001-8560-8397 |
| Yong Kong | | 203-785-7662 | 0000-0002-2881-5274 |
| Morgan Roos | | 608-442-6100 | 0000-0003-0325-1334 |
| Christina Middle | | 608-442-6100 | |
| Hanqiao Zhang | | 626-395-3464 | 0000-0001-7394-8781 |
| Matt Thomson | | 626-395-8782 | |
| Keith Booher | | 888-882-9682 | 0009-0006-1068-4057 |
| Scott Kuersten | | 608-442-6100 | 0009-0007-5058-4920 |
| **Corresponding Authors** | | | |
| Akiko Iwasaki | | 203-785-7662 | 0000-0002-7824-9856 |
| Rustem F Ismagilov | | 626-395-8130 | 0000-0002-3680-4399 |
